# Polygenic Risk Score Improves the Accuracy of a Clinical Risk Score for Coronary Artery Disease

**DOI:** 10.1101/2022.06.02.22275933

**Authors:** Austin King, Lang Wu, Hong-Wen Deng, Hui Shen, Chong Wu

## Abstract

**Background:** The value of polygenic risk scores (PRS) towards improving guideline-recommended clinical risk models for coronary artery disease (CAD) prediction is controversial. Here we examine whether an integrated polygenic risk score improves prediction of CAD beyond pooled cohort equations.

**Methods:** An observation study of 291,305 unrelated White British UK Biobank participants enrolled from 2006 to 2010 was conducted. A case-control sample of 9,499 prevalent CAD cases and an equal number of randomly selected controls was used for tuning and integrating of the polygenic risk scores. A separate cohort of 272,307 individuals (with follow-up to 2020) was used to examine the risk prediction performance of pooled cohort equations, integrated polygenic risk score, and PRS-enhanced pooled cohort equation for incident CAD cases. Performance of each model was analyzed by discrimination and risk reclassification using a 7.5% threshold.

**Results:** In the cohort of 272,307 individuals (mean age, 56.7 years) used to analyze predictive accuracy, there were 7,036 incident CAD cases over a 12-year follow-up period. Model discrimination was tested for integrated polygenic risk score, pooled cohort equation, and PRS-enhanced pooled cohort equation with reported C-statistics of 0.640 (95% CI, 0.634-0.646), 0.718 (95% CI, 0.713-0.723), and 0.753 (95% CI, 0.748-0.758), respectively. Risk reclassification for the addition of the integrated polygenic risk score to the pooled cohort equation at a 7.5% risk threshold resulted in a net reclassification improvement of 0.117 (95% CI, 0.102 to 0.129) for cases and -0.023 (95% CI, -0.025 to -0.022) for noncases [overall: 0.093 (95% CI, 0.08 to 0.104)]. For incident CAD cases, this represented 14.2% correctly reclassified to the higher-risk category and 2.6% incorrectly reclassified to the lower-risk category.

**Conclusions and Relevance:** Addition of the integrated polygenic risk score for CAD to the pooled cohort questions improves the predictive accuracy for incident CAD and clinical risk classification in the White British from the UK biobank. These findings suggest that an integrated polygenic risk score may enhance CAD risk prediction and screening in the White British population.

## Background

Cardiovascular disease (CVD) is a major cause of death worldwide.^1^ Risk estimates for CVD have become particularly important for disease prevention and clinical practice.^2,3,4,5^ Current guidelines from the American College of Cardiology and American Heart Association suggest lipid-lowering treatments for individuals with greater than a 7.5% 10-year absolute risk of developing CVD based on pooled cohort equations (PCE).^6^ Because of the central role of accurate risk estimates in CVD prevention, improving accuracy beyond those already used in clinical practice like PCE, could save lives by better identifying high risk individuals.

Substantial advancements have been made over the past decades in identifying genetic variants associated with coronary artery disease (CAD).^7,8,9,10^ Recent advances in polygenic risk scores (PRSs) have sparked a great interest in enhancing disease risk prediction by using the information on millions of variants across the genome.^11,12,13,14^ However, population health utility of PRSs in CAD risk prediction is controversial. Several studies have shown that PRSs can improve risk prediction accuracy for incident and prevalent CAD cases compared with individual conventional risk factors^15,16^ and combing risk prediction models (like PCE) with PRS improves the performance in terms of net reclassification improvement.^17^ On the other hand, several studies^18,19^ integrating PRSs into PCE to assess possible clinical utility have concluded that the current benefits of incorporating PRSs were minimal (although statistically significant) and were not considered clinically significant to warrant their use over current clinical used prediction models. In this manuscript, we investigate why different studies have reached different and controversial conclusions. Specifically, we analyzed UK Biobank data to test the hypothesis that integrated PRSs leveraging multiple newly developed PRS methods, and several genome-wide association study (GWAS) datasets, can improve risk prediction for CAD over the widely used PCE and thus provide improved clinical utility in European populations.^9,20,21,22,23,24,25^ Furthermore, in secondary analysis, we extended our integrated method to analyze its predictive performance in non-European populations.

## Methods

### Study Populations

Our study utilized the UK Biobank which includes 502,536 participants ranging in age from 40 to 69 at baseline recruitment.^26^ Biomarker data were collected from stored serum and red blood cells, details of which are described elsewhere.^27^ Ethical approval for the UK Biobank study was obtained from the National Health Service’s National Research Ethics Service North West (11/NW/0382). The current research project (application number 48240) was approved by UK Biobank. Our study design is outlined in **Figure 1**.

**Figure 1.**
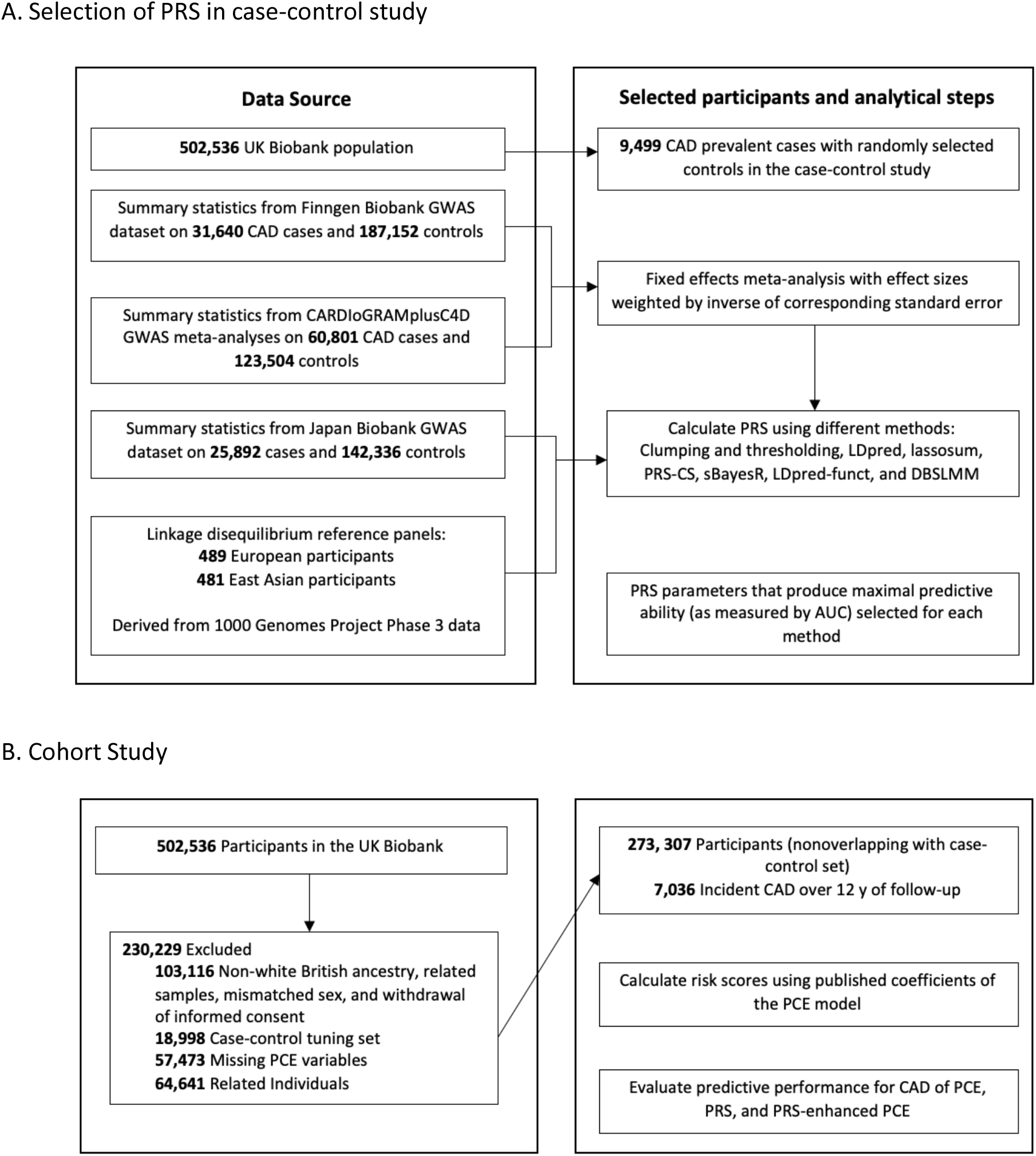
Study Design and Flowchart for Coronary Artery Disease (CAD) To select the parameters for each method with the best discrimination based on area under the curve (AUC), clumping and thresholding, LDpred, lassosum, PRS-CS, sBayesR, LDpred-funct, and DBSLMM were used to calculate polygenic risk scores (PRS) on the case-control set consisting of prevalent cases. For these calculations, summary data for three genome-wide association studies (GWAS) on CAD (CARDIoGRAM-plusC4D, Finngen Biobank, Japan Biobank) that excluded UK Biobank and data on linkage disequilibrium were used. The calculated PRS were applied to a nonoverlapping set of participants from the UK Biobank with no preexisting CAD, aged 40 to 69 at baseline, and who were followed up for incident CAD events. In this population, the pooled cohort equations (PCE) model was calculated and different models (PRS, PCE, PRS-enhanced PCE) were compared in terms of their predictive accuracy based on discrimination, calibration, and reclassification metrics.

The primary endpoint for our study was CAD, for which several large GWAS results are available.^8,9,25^ We limited our primary investigation to unrelated White British individuals (as defined by UK Biobank data-field 22006) to reduce the influence of population heterogeneity and related samples; unrelated individuals were obtained by only keeping individuals with no relative 3^rd^ degree or closer.^28^ We further excluded outliers for heterozygosity or genotype missing rates (0.2> missing rate). Participants with inconsistent reported and genotypic inferred sex and withdrawn consent were likewise removed.

In secondary analysis, we focused on African ancestry participants in the UK Biobank. Following others,^29,30^ we used imputed data released by the UK Biobank to determine continental ancestry (African (AFR), East Asian (EAS), European (EUR), South Asian (SAS)) and projected participants onto genetic principal components calculated in the 1000 Genome Project (N= 2000: AFR = 504; EAS = 504; EUR = 503; SAS = 489). We excluded populations identified as African Caribbeans in Barbados (ACB) and Americans of African Ancestry in SW (ASW) from the AFR population and all individuals of American ancestry (AMR) due to their complex admixture patterns. Participants were assigned to ancestries based on likelihoods calculated from their first 5 principal components. Samples were assigned via random forest to an ancestry when their likelihood for a given ancestry was > 0.3. If two ancestries exceeded 0.3, we assigned ancestries as: AFR over EUR, SAS over EUR, and EUR over EAS. Participants were excluded if no likelihood was > 0.3 or if 3 ancestry groups were > 0.3 (n = 8). The same quality control used in primary analysis was then applied to the resulting AFR ancestry population.

The study population was divided into (1) a case-control study (tuning dataset) established from prevalent CAD cases (see Cardiovascular Outcome Definitions Subsection for details) and randomly selected controls and (2) an independent prospective cohort study (testing dataset) of participants with no history of CAD at baseline recruitment. The tuning dataset was used for building risk prediction models and the testing dataset was used for unbiasedly evaluating their performance. Of note, there were no overlapping participants between these two datasets, ensuring the testing results were valid.

### Definition of Risk Score Variables

The updated pooled cohort equation (PCE) model, a clinically used risk prediction model, was used as our baseline. We matched variables available in the cohort to the predictors of the updated PCE,^3^ including information on age, sex, race and ethnicity, smoking status, total and HDL cholesterol, systolic blood pressure, diabetes, and the use of lipid lowering and blood pressure lowering medications. Definitions for Type 1 and Type 2 diabetes, blood pressure lowering and lipid lowering medications use as well as categorization of smoking status were defined based on UK-recommended QRISK3 scores.^31,32^ Details of variable definitions and protocol for handling missing values are relegated to the eMethods section of Supplementary.

### Cardiovascular Outcome Definitions

The UK Biobank data have been linked to Hospital Episode Statistics (HES) and national death and cancer registries. HES records diagnosis information via International Classification of Diseases (ICD)-9^th^ and 10^th^ Revisions and codes operative procedures via OPCS-4. Death registries include death date and both primary and secondary causes of death coded in ICD-10. We defined CAD by combining HES, death registries, operation codes,^31,32^ as well as related self-reported diagnoses and previous procedure codes (Supplementary Tables 1 and 2). Following others,^18^ CAD was defined as myocardial infarction, including related sequelae.

The date of event was determined via recorded episode date, admission date, or operation date indicated in the hospital statistics. For participants with multiple CAD event dates, the earliest recorded date were used as the dates of event. Age of event was determined by self-reported age and calculated age based on the date of event; when both ages were available, the smaller value was used.^15^ Prevalent cases at baseline were defined as individuals with age of event earlier than age at UK Biobank enrollment time. Follow-up time was calculated as the number of days from assessment date until the event of interest (CAD event), a competing cause of death, or censorship date (2020/12/31) occurred.

### **Polygenic Risk Scores** (PRS)

Information on genotyping and imputation has been described in detail elsewhere.^27,33^ Standard quality-control procedures were applied to the imputed UK Biobank genotype data. Briefly, we restricted our analyses to autosomal genetic variants, kept variants with imputation information score (INFO) score > 0.3, minor allele frequency > 1%, Hardy-Weinberg equilibrium P > 10^−10^, and genotype missing rate < 10%. We further removed variants with ambiguous strands (A/T or C/G).

PRS for CAD were derived as weighted sums of risk alleles using 3 CAD GWAS datasets (CARDIo-GRAMplusC4D, FinnGen Biobank, Japan Biobank) that had no overlap with the present UK Biobank study (**Figure 1**).^8,9,25^ The 3 GWAS datasets were filtered to only include SNPs present in the imputed UK Biobank data. For all datasets, we aligned β and allele frequencies to the hg19 alternate allele. First, we performed a fixed-effect meta-analysis focused on GWAS datasets with subjects of European ancestry, specifically the CARDIoGRAMplusC4D and FinnGen datasets, using METAL.^34^ Second, the PRSs were calculated by using either Japan Biobank data or combined European data and their corresponding population-specific 1000 Genome Project constructed LD reference panels.

Tuning of the PRS was implemented using seven methods: (1) clumping and thresholding using PRSice-2 software (version 2.3.3),^35^ (2) LDpred,^20^ (3) lassosum,^36^ (4) PRS-CS,^21^ (5) sBayesR,^22^ (6) LDpred-funct,^23^ and (7) DBSLMM.^24^ Detailed information on each PRS method and their associated parameters are described in the eMethods section of Supplementary. All methods utilized were adjusted for genotype measurement batch and the first five genetic principal components calculated by the UK Biobank. Since different PRS methods and datasets may capture different information, we constructed the integrated PRS by 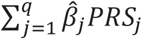, where 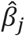 is the estimated coefficient of *PRS*_*j*_ in the logistic regression using the tuning dataset and *PRS*_*j*_ is the j^th^ PRS.^37^ Selection of PRS methods for the integrated model was determined based on are under the curve (AUC) results from the tuning dataset. Methods with the largest AUC improvement over the PCE model were selected and analyzed in the testing dataset until the inclusion of additional PRS methods failed to improve the predictive performance of the integrated model. We assessed the performance of the integrated model against the individual PRS methods in the testing dataset as well as models combining the European meta-analysis data and Japan Biobank data.

### Statistical Analysis

Participants were excluded from the study for multiple factors, including missing genetic data, mismatches in reported and genotypic sex, withdrawal of informed consent, and missing predictor values. Using previously published baseline coefficients for each predictor variable and baseline hazard, we calculated the updated pooled cohort equation scores (PCE).^3^ We examined several models as defined in previous studies^18,19^: (1) PCE; (2) (integrated) PRS for CAD; and (3) PCE and (integrated) PRS. We performed Cox proportional hazard regression using follow-up time as the time variable in the testing data. As a sensitivity analysis, all models were reexamined after removing participants that reported taking lipid-lowering medications at baseline of the UK Biobank study.

We examined the discrimination of each model via Harrell’s C-statistic and its 95% confidence interval. ^38,39,40^ In brief, the C-statistic is a measure of the discriminatory power of a risk prediction model, with values ranging from 0.5 (no discrimination) to 1.0 (perfect discrimination). Calibration and recalibration of the baseline models were graphically assessed by comparing observed probabilities via Kaplan-Meier estimates to the mean predicted probability within tenths of the predicted probabilities. During recalibration, the baseline survival function was estimated in the testing cohort and combined with predicted hazard ratios from the validation dataset in a Cox model to obtain recalibrated predicted probabilities.^3,18^ We assessed the recalibration results via the calibration slope and Greenwood-Nam-D’Agostino test.^41^

We evaluated risk prediction accuracy using the net reclassification improvement (NRI)^42^ at a threshold of 7.5% (clinically used in the United States), continuous NRI, and associated integrated discrimination improvement (IDI).^43^ These metrics quantify how well a new model (PCE plus PRS) reclassifies individuals compared to an old model (PCE); a brief explanation of these metrics can be found in the eMethods section of the Supplementary.

Statistical analyses were conducted in R software, version 4.0.0 (R Project for Statistical Computing).^44^ Anaconda, version 3.8.3, was also used for PRS methods that utilized Python programming language.^45^

## Results

Following removal of participants with missing data and selecting for only unrelated white British participants, the UK Biobank dataset contained 291,305 participants which were subsequently divided into case-control and cohort study datasets (**Figure 1**). The case-control study contained 9,499 prevalent CAD cases and an equal number of controls used for tuning of the PRS methods. The independent cohort study was comprised of 272,307 individuals (mean age: 56.7) with 7,036 incident cases. Participants with CAD at baseline were not included in the cohort study population. The cohort study had a median follow-up time of 12.33 years (interquartile range, 1.42), while incident CAD cases had a median follow-up time of 5.02 years (interquartile range, 4.07). Baseline characteristics (such as age, smoking status, cholesterol, and systolic blood pressure) were similar for participants included in the cohort analysis and excluded due to missing covariates (Supplementary Tables 3A-3C).

For the case-control study, each PRS method for CAD was performed across multiple parameter settings to determine optimal values that would be combined for the cohort study. We classified the “optimal” parameter values as those achieving the highest AUC values for that individual method. Specific details on each method’s tuning parameters and individual AUC values were provided in Supplementary Tables 4A and 4B for the European meta-analysis (EUR) and Japan Biobank (Japan) datasets. We combined the PRS for CAD based on the combination of the three GWAS datasets. As expected, because the combined EUR + Japan methods fully utilized all three GWAS datasets and several complementary PRS methods, it achieved the highest AUC [0.641 (95% CI, 0.635-0.648)] and thus we focused on this PRS (denoted by integrated CAD PRS or simply PRS) for the remaining analysis. The maximal integrated CAD PRS model for this study was determined to include the EUR and Japan derived clumping and thresholding, LDpred, lassosum, PRS-CS, and LDpred-funct methods. During this step, we evaluated the PRS methods for collinearity concerns and determined the different methods tended to not be highly correlated (Supplementary Figure 1).

In the cohort analysis, following selection of white British participants, as well as excluding individuals with missing data, and selecting the case-control subjects, 272,307 participants were used. The discrimination of the integrated CAD PRS remained similar as that in tuning case-control study; the C-statistic for the integrated CAD PRS was 0.640 (95% CI, 0.634 -0.646) (**Table 1**). The discrimination of the PCE (C-statistics, 0.718 [95% CI, 0.713-0.723]) was higher than the integrated CAD PRS. Addition of individual PRSs to the PCE resulted in improved discrimination of the model with PRS-CS applied to the European meta-analysis showing the highest discrimination (C-statistics, 0.749 [95% CI, 0.744-0.754]) (Supplementary Table 5A-5C). We observed the most significant improvement in discrimination when the integrated CAD PRS were added to the PCE, showing a C-statistic increase to 0.753 (95% CI, 0.748-0.758), an associated change over the PCE alone of 0.035 (95% CI, 0.03 – 0.04) (**Table 1** and **Figure 2**). We further stratified the population by age group (younger and older than 55 years of age) and sex (men and women) separately and observed higher discrimination in women than men and higher discrimination in the younger age group than in the older age group (**Table 1**). Participants that were not receiving lipid-lowering medication at baseline were also examined and demonstrated similar discrimination performance (**Table 1**).

**Table 1.** C-Statistics for Coronary Artery Disease for Full Population and Stratified by Sex and Age Group (Younger and Older than 55 Years of Age) ^1,2^.

| C-Statistic (95% CI) |  |  |  |  |  |  |
| --- | --- | --- | --- | --- | --- | --- |
|  | All | Men | Women | Participants<br>Aged <55 y | Participants<br>Aged ≥ 55 y | Participants<br>Not Receiving<br>Lipid-Lowering<br>Treatment at<br>Baseline |
| **A. White British Ancestry** |  |  |  |  |  |  |
| Participants, No. | 272307 | 124155 | 148152 | 102330 | 169977 | 235172 |
| Cases, No. | 7036 | 5093 | 1943 | 1276 | 5760 | 5091 |
| Polygenic risk<br>score | .64 (.634-.646) | .643 (.636-.651) | .641 (.629-.654) | .69 (.626-.705) | .632 (.625-.639) | .646 (.638-.653) |
| Pooled cohort<br>equation | .718 (.713-.723) | .663 (.656-.67) | .706 (.695-.717) | .749 (.736-.761) | .665 (.658-.671) | .73 (.724-.737) |
| Polygenic risk<br>score + pooled<br>cohort equation | .753 (.748-.758) | .714 (.708-.721) | .741 (.73-.751) | .793 (.781-.806) | .705 (.699-.712) | .766 (.76-.772) |
| **B. African Ancestry** |  |  |  |  |  |  |
| Participants, No. | 6753 | 2901 | 3852 | 4528 | 2225 | 5896 |
| Cases, No. | 88 | 46 | 42 | 42 | 46 | 63 |
| Polygenic risk<br>score | .542 (.485-.6) | .574 (.494-.654) | .6 (.511-.634) | .548 (.46-.637) | .543 (.462-.624) | .534 (.464-.604) |
| Pooled cohort<br>equation | .714 (.659-.769) | .674 (.595-.753) | .734 (.653-.815) | .657 (.572-.742) | .721 (.656-.787) | .698 (.628-.768) |
| Polygenic risk<br>score + pooled<br>cohort equation | .716 (.665-.768) | .695 (.622-.768) | .732 (.654-.81) | .679 (.597-.761) | .696 (.629-.763) | .707 (.64-.774) |

1. Cox proportional hazard models for CAD using recalibrated polygenic risk score, pooled cohort equations, and both combined models. 2. C-Statistics shown for combined European meta-analysis + Japan Biobank PRS methods. Results are presented for both White British and African ancestry populations.

**Figure 2.**
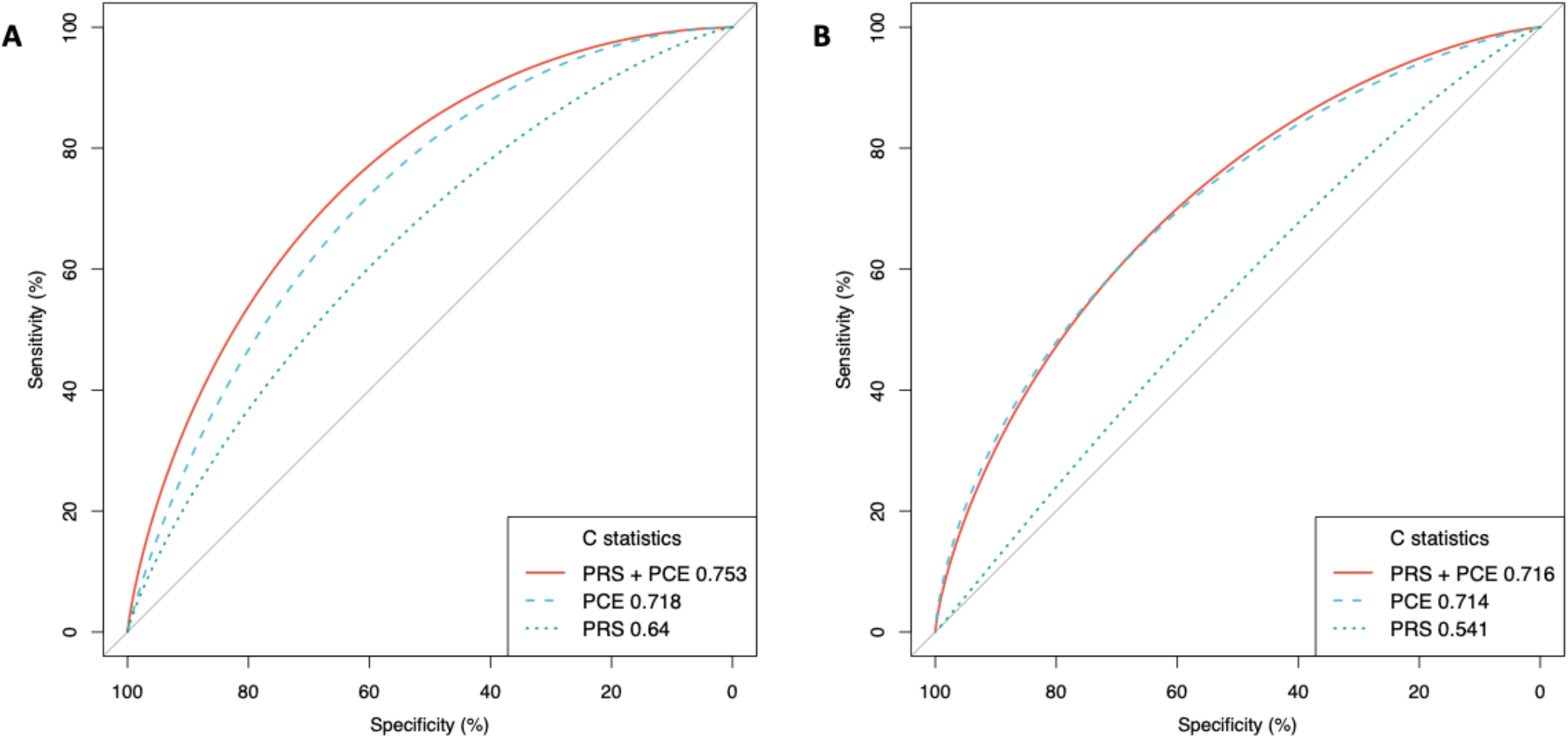
Receiver Operator Characteristic Curves and C-Statistics for Different Models in Cohort Analyses of White British and African Ancestry Populations. PCE indicates pooled cohort equation; PRS indicates integrated polygenic risk score. A) is the White British population of 272,307 individuals over a mean 12 years of follow-up with 7036 incident CAD cases and B) is the African ancestry population of 6,753 individuals over a mean 13 years of follow-up with 88 incident CAD cases.

When evaluating model performance, we compared observed and predicted cumulative incidences of CAD events across each tenth of predicted risk and determined the addition of our integrated PRS method to the baseline model overestimated risk. Following others,^17,18^ we recalibrated the model by fitting predicted log-HRs as covariates in the model, resulting in considerable improvement in model calibration (Supplementary Figure 2).

We investigated the potential of the PRS-enhanced PCE model in the risk assessment of CAD. We found that an individual’s integrated CAD PRS were generally uncorrelated (Pearson correlation coefficient r, 0.01) with their PCE, which partially explains why adding integrated CAD PRS to PCE model (denoted by PRS-enhanced PCE) improves the discrimination power. We evaluated the hazard ratios HR via a Cox regression. The PCE model had an adjusted HR of 1.653 (95% CI: 1.628-1.679) per standard deviation increase (*P* < .001) while the PRS-enhanced PCE model reported an adjusted HR of 1.77 (95% CI: 1.745 - 1.796) per standard deviation increase of PRS (*P* < .001). The PRS-enhanced PCE model further improves the discrimination power of PCE model (**Figure 3**). For example, in the PRS-enhanced PCE model, there was a 7.77-fold (95% CI: 7.61- to 7.92-fold) risk of CAD for individuals in the top quintile compared to those in the bottom quintile. The PCE model, in comparison, reported a 5.29-fold (95% CI: 5.21- to 5.39-fold) risk of CAD between the top and bottom quintiles.

**Figure 3.**
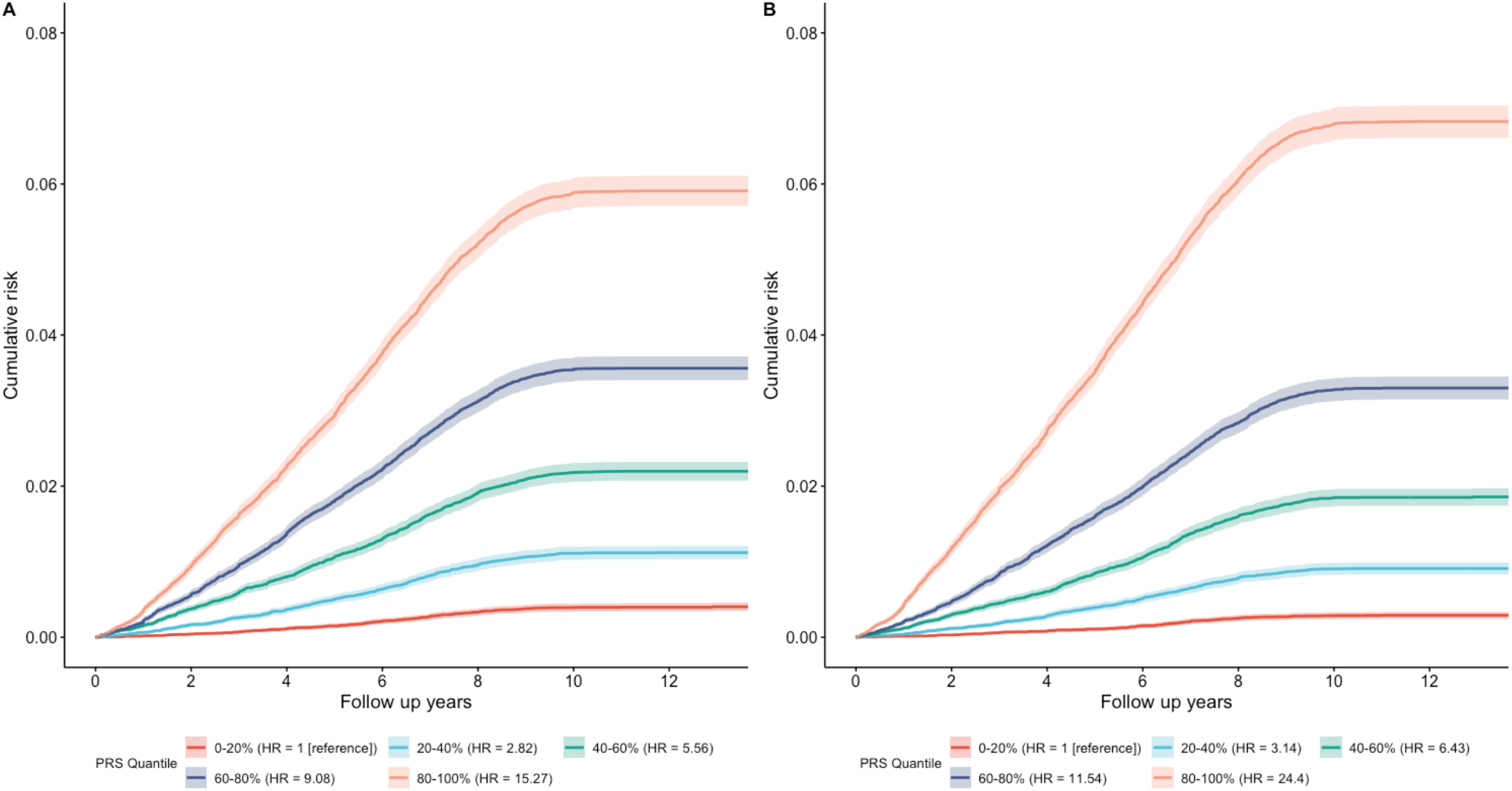
Cumulative Absolute Risk of Developing CAD. Cumulative absolute risk of developing CAD by quintiles of the overall polygenic score in **A)** the PCE model and **B)** the PRS-enhanced PCE model. The shaded portions correspond to the 95% confidence interval.

After adding PRS for CAD to the PCE model, predicted risk changed by greater than 1% for 35.5% of participants while changing by 5% or greater for 1.9% of participants (**Figure 4A**). There were 7,005 incident CAD cases and 256,072 noncases at the 10-year follow-up; 9,230 individuals were censored due to lack of disease or follow-up at 10 years. At the suggested 7.5% risk threshold, 992 of 7,005 cases (14.2%) were correctly reclassified to the higher-risk category and 182 of 7,005 cases (2.6%) were incorrectly moved to the lower-risk category. For non-case participants, 3,443 of 256,072 (1.3%) were correctly moved down to the lower-risk category while 9,331 of 256,072 (3.6%) were incorrectly moved up to the high-risk category (**Figure 4B**).

**Figure 4.**
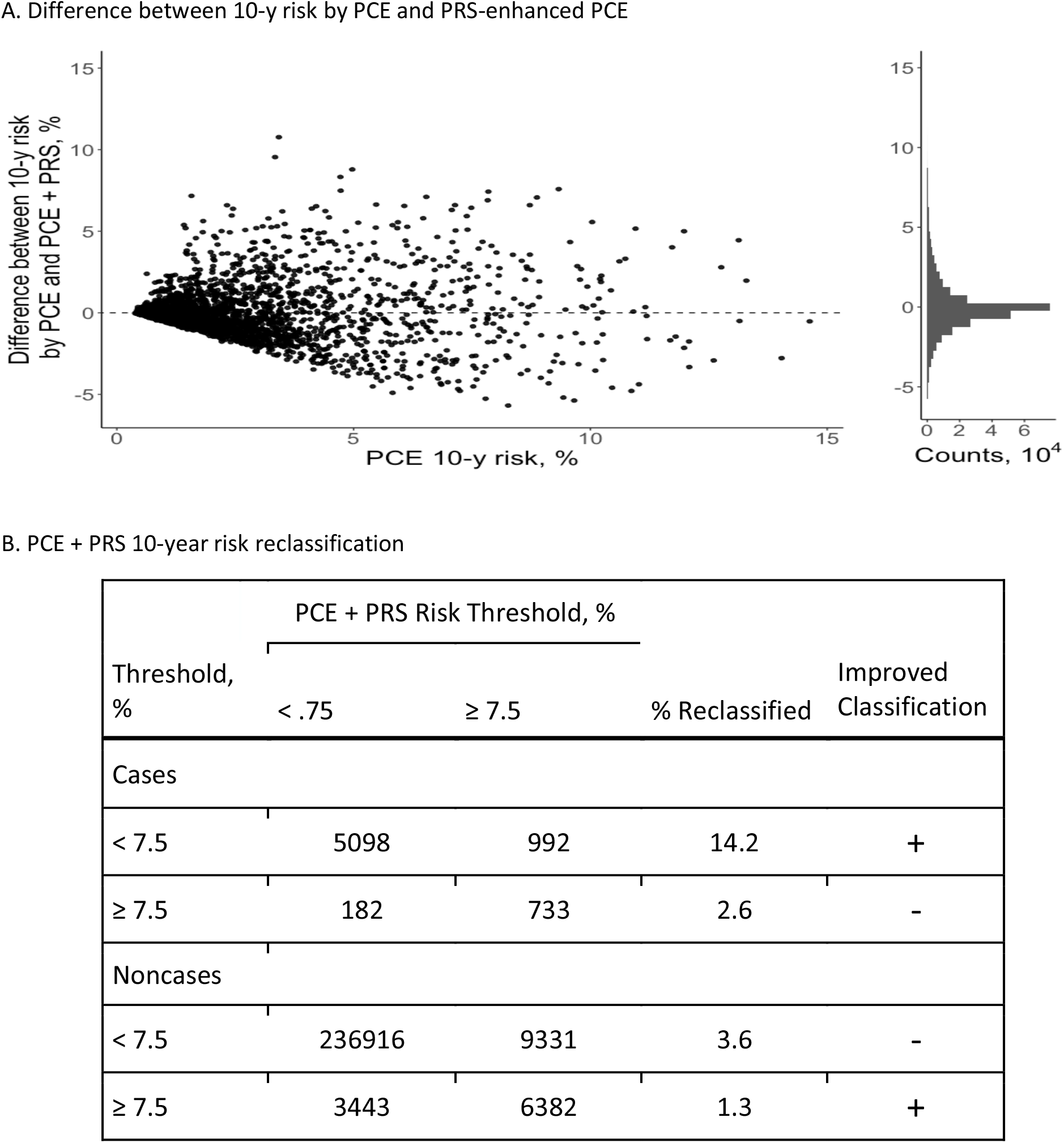
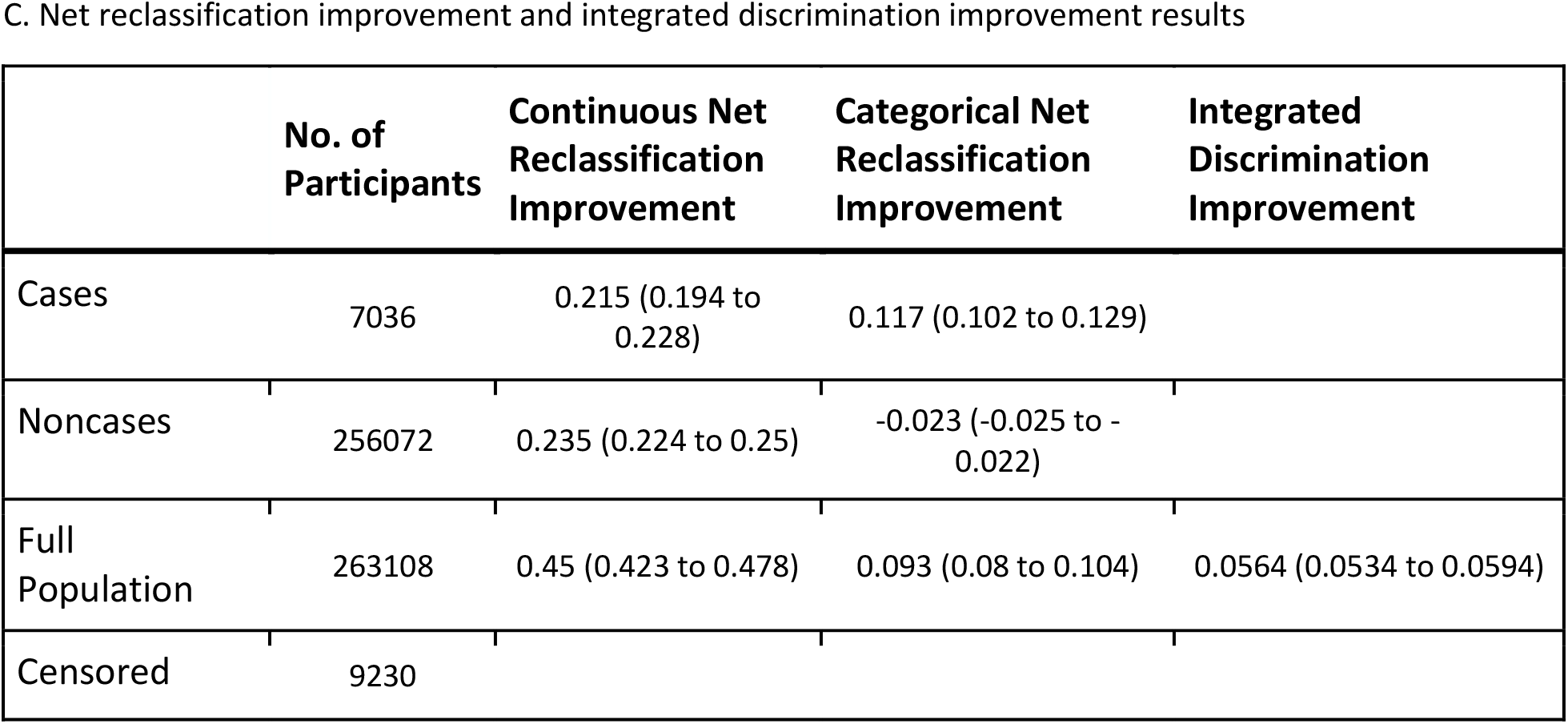
Change in Predicted Probabilities and Risk Reclassification. A, Change in the predicted probabilities of the recalibrated pooled cohort equations (PCE) model after the addition of polygenic risk scores (PRS) for CAD. The x-axis shows the predicted probability from the baseline PCE model. The y-axis is the difference in 10-year risk probabilities of a CAD event between the PRS-enhanced model and the baseline PCE model. The scatterplot has a random draw of 1% of the participants shown. The histogram x- and y-axes are based on the full population. B, Reclassification table of predicted probabilities by PCE and PRS-enhanced PCE models at 7.5% threshold. Rows indicating an improved classification with the PRS-enhanced PCE model are marked by a plus sign while rows indicating a deteriorated classification are marked by a minus sign. C, Table of net reclassification improvement (NRI) and integrated discrimination improvement (IDI). NRI^a^ is defined in the continuous case as the sum of proportions of cases and noncases with improved combined score minus the sum of proportions with deteriorated combined score. In the categorical case, NRI is defined by changed at a 7.5% threshold predicted probability. A positive NRI indicates a better combined score overall. IDI^b^ measures the difference of average probabilities of an event in cases and noncases. A larger IDI indicates more discrimination in the combined score. ^a^ NRI = P(up|case) - P(down|case) - P(up|noncase) + P(down|noncase) ^b^ IDI = P_PCE+PRS_(case) - P_PCE+PRS_(noncase) - P_PCE_(case) + P_PCE_(noncase)

When comparing integrated PRS for CAD model to the PCE model, the NRI for cases was 11.7% (95% CI, 10.2% to 12.9%) and –2.3% (95% CI, -2.5% to –2.2%) for noncases (**Figure 4C**). Following the addition of the integrated CAD PRS to PCE, the IDI metric indicated an increase in risk difference between cases and noncases of 0.056 (95% CI, 0.053 to 0.059) (**Figure 4C**). Stratification by sex indicated higher NRI improvement in men over women; stratification by age group saw similar overall NRI improvement (Supplementary Table 6).

### Secondary Analyses

There were 6,971 participants in the AFR ancestry population that were divided into case-control and cohort datasets. The case-control dataset consisted of 109 prevalent CAD cases and an equal number of controls. The cohort population was composed of 6,753 participants (median follow-up: 12.75, interquartile range: 1.25) in which 88 incident CAD cases were observed (median follow-up: 5.97, interquartile range: 3.3). Baseline characteristics are presented in Supplementary Tables 3D-3F.

In case-control analysis, the optimized integrated CAD PRS model that achieved the highest AUC (0.717 [95% CI, 0.644-0.769]) was determined to include the EUR clumping and thresholding, LDpred, PRS-CS, and LDpred-funct methods as well as the Japan LDpred, PRS-CS, and sBayesR methods. In cohort analysis, the integrated CAD PRS C-statistic was 0.542 (95% CI, 0.485-0.6) (**Figure 1**). Discrimination of the PCE model (0.714 [95% CI, 0.659-0.769]) outperformed the integrated CAD PRS. In contrast to the White British population, the incremental value of the addition of the integrated CAD PRS to the PCE model was minimal (increase in C-statistic, .002 [95% CI, 0.006 to -0.001]) (**Table 1**). We further stratified by gender and age and observed higher discrimination in women and in the older age group, however, we noticed slightly greater improvement in discrimination with the addition of our integrated CAD PRS in both men and the younger age group. Participants not on lipid-lowering medication at baseline saw slightly higher, but still minimal discrimination improvement than the full population (**Table 1**). NRI and IDI metrics were likewise minimal and incrementally smaller than in the White British population (Supplementary Table 7).

## Discussion

In our analysis, the addition of genetic information to the PCE clinical risk score was associated with a moderate improvement in predictive accuracy for CAD. Addition of PRS to the baseline PCE model resulted in a 3.5% improvement in model concordance as well as a 9.3% net reclassification improvement (NRI) of incident CAD cases and noncases over the baseline PCE model at a 7.5% risk threshold. In comparison, the integrated risk tool^17^ and Elliott et al.^18^ achieved 2.7% (in European population) and 4.0% (in all UK Biobank subjects) improvement in terms of NRI, respectively. While both studies improve the performance by integrating PRS into PCE, they reached different conclusions regarding its clinical utility, highlighting the importance of building a more powerful and accurate risk prediction model.

Our studies are innovative and are different from existing studies evaluating the clinical utility of adding PRS over existing clinical risk models^18,19,46,47^ in the following aspects. While matching our definition of CAD to that of a previous study performed with the UK Biobank,^18^ we were able to take advantage of more recent incident CAD data. We also utilized a stricter definition for our target population in the UK Biobank data as opposed to the entire UK Biobank data, which contain individuals of diverse ancestry. Recent studies have shown population-specific bias and limited use of specific PRS methods when used on non-European populations.^48,49^ We also used three distinct GWAS datasets to build the PRS and integrated results from several advanced and more recent PRS methods,^21,22,23,24^ improving the discrimination power of our integrated CAD PRS.

We found that integrating PRS to baseline PCE model resulted in significant continuous and categorical NRI. Categorical NRI for incident cases was 11.7% and -2.3% for noncases. Our model greatly improved reclassification for cases over previous studies,^17,18,19^ but resulted in more misclassification in non-case individuals. This difference in performance for noncases may be due in part to model specifications and cohort selection. In contrast to Moseley et al,^19^ in which the 2013 PCE model was used, we utilized the updated 2018 PCE as our baseline. The 2013 model was noted to overestimate risk across all risk groups, prompting the development of the updated PCE model.^3^ We also used a younger cohort compared to the two cohorts in Moseley et al (mean age 56.7 years compared to 62.9 and 61.8, respectively). As noted, we included only White British ancestry in our primary cohort. The inclusion of other ethnicities in the cohort may significantly decrease the discrimination power of the PRS constructed. This is shown in our secondary analysis of African ancestry, where the PRS results based on a European ancestry GWAS dataset vastly underperformed compared to the White British population (C-statistics 0.715 vs 0.752, respectively) (Supplementary Table 5).

Our results suggest an association between predictive accuracy of PRS and incident CAD events that varies based on age and sex. Men showed significantly higher C-statistic improvement than women (0.051 vs 0.035) in the PRS-enhanced PCE model over the baseline PCE model. This is complemented by an 11.6% overall categorical NRI improvement in men compared to 3.6% in women (Supplementary Table 6). Recent studies using PRS in the UK Biobank demonstrated comparable results with higher risks for incident CAD in men than women.^15,47,50^ The improved performance in men may be attributed to overrepresentation of male CAD cases in the case-control and cohort studies. The use of sex-specific data may lead to improved prediction accuracy of PRS.

Our results also suggest a genetic component to early-onset cases of CAD and a possible application of PRS in identifying individuals at heightened risk of these cases, as the predictive accuracy of incident CAD cases was higher in participants < 55 years of age. The observed C-statistic for the integrated PRS-enhanced PCE model was 0.793 compared to 0.705 observed in the ≥ 55 age group. This observation supports two recent studies that found high risk score predictions in genetic variants strongly associated with early-onset CAD (<40 years old) as well as improved risk classification of early-onset CAD to higher-risk categories that were not classified as such by PCE.^9,51^

There are limitations in our study. First, our study was conducted in the UK Biobank and is, therefore, limited by the characteristics of the cohort. The UK Biobank cohort is composed of primarily European ancestries (further restricted to White British ancestry in this study) and limited to an age range of 40 to 69 years, restricting its application to other ancestries and age groups. In addition, participants in the UK Biobank assessment tend to be healthier and more well-off compared to the general UK population,^52^ and thus population-level CAD risk may be underestimated in our study. In secondary analysis, the limited genetic diversity of the UK Biobank cohort is apparent and resulted in significantly smaller tuning and testing. The extent to which our results can be applied to larger non-European ancestries, in particular African ancestry, warrants further investigation. These results also highlight the urgency of developing novel cross-ancestry PRS methods^10,17,53,54,55^ and using more diverse cohorts to construct PRSs.^17^ In addition, as the case-control and cohort analyses are derived from the same study, more broad generalizability of the results requires further investigation. Second, this study included PRS for low frequency and common genetic variants (MAF ≥ 1%) and did not examine the predictive accuracy of rare variants known to affect CAD risk. Third, the algorithm for selection of CAD cases utilizes self-report, death, and hospital inpatient data for the definition of prevalent and incident CAD cases. As such, misclassification of cases is possible. Fourth, tuning of each PRS method in the case-control study used prevalent CAD cases, which could introduce survival bias. However, simulation studies have demonstrated a limited effect of survival bias on estimated genetic effects of event risks.^56^ Fifth, participants with at least 1 missing predictor value were excluded from the study. Excluded participants were not considerably different demographically from those included and thus the missing data are unlikely to have a significant effect on the reported estimates.

## Conclusions

Addition of the integrated CAD PRS to the PCE resulted in a statistically significant improvement in predictive accuracy for incident CAD, especially in individuals under the age of 55 years old in White British population. It was also associated with moderate improvement in risk reclassification across all subgroups. However, the benefits of adding integrated CAD PRS to the PCE are minimal for African population. In summary, the inclusion of genetic information to the pooled cohort equation can help improve clinical risk classification and demonstrates the potential for genetic screening in early life to improve clinical risk prediction in White British population.

## Data Availability

UK Biobank data used in this study were available upon UK Biobank approval (https://www.ukbiobank.ac.uk, application number 48240). The summary statistics of genome-wide association studies (GWAS) of FinnGen Biobank can be obtained from https://www.finngen.fi/en/access_results upon registration, CARDIoGRAMplusC4D GWAS data can be directly downloaded at
http://www.cardiogramplusc4d.org/data-downloads/, Japan Biobank GWAS data can be downloaded at https://humandbs.biosciencedbc.jp/en/hum0014-v22#42diseases.
1000 Genomes phase 3 reference panel can be obtained at
https://www.internationalgenome.org/data-portal/data-collection/phase-3. The code can be downloaded from https://github.com/ChongWuLab/PolygenicRiskScore_CAD.

http://www.cardiogramplusc4d.org/data-downloads/

https://humandbs.biosciencedbc.jp/en/hum0014-v22#42diseases

https://www.internationalgenome.org/data-portal/data-collection/phase-3

https://github.com/ChongWuLab/PolygenicRiskScore_CAD

## Abbreviations

(CVD): Cardiovascular disease;
(PCE): pooled cohort equations;
(CAD): coronary artery disease;
(PRS): polygenic risk score;
(GWAS): genome-wide association study;
(AFR): African;
(EAS): East Asian;
(EUR): European;
(SAS): South Asian;
(HES): hospital episode statistics;
(AUC): area under the curve;
(NRI): net reclassification improvement;
(IDI): integrated discrimination improvement;
(CI): confidence interval

## References

1. GBD 20019 Disease and Injuries Collaborators. Global burden of 369 diseases and injuries in 204 countries and territories, 1990-2019: a systematic analysis for the Global Burden of Disease Study 2019. Lancet. 2020; 396(10258):1204–1222.

2. Damen JA, Hooft L, Schuit E, et al. Prediction models for cardiovascular disease risk in the general population: systematic review. BMJ. 2016;353:i2416.

3. Yadlowsky S, Hayward RA, Sussman JB, McClelland RL, Min YI, Basu S. Clinical implications of revised pooled cohort equations for estimating atherosclerotic cardiovascular disease risk. Ann Intern Med. 2018;169(1):20–29.

4. Conroy R, Pyörälä K, Fitzgerald A, et al. Estimation of ten-year risk of fatal cardiovascular disease in Europe: The SCORE project. European Heart Journal. 2003;24(11):987–1003.

5. D’Agostino R, Vasan R, Pencina M, et al. General Cardiovascular Risk Profile for Use in Primary Care. Circulation. 2008;117(6):743–753.

6. Arnett DK, Blumenthal RS, Albert MA, et al. 2019 ACC/AHA guideline on the primary prevention of cardiovascular disease: a report of the American College of Cardiology/American Heart Association Task Force on Clinical Practice Guidelines. J Am Coll Cardiol. 2019;74(10):e177–e232.

7. Musunuru K, Kathiresan S. Genetics of common, complex coronary artery disease. Cell. 2019;177(1):132–145.

8. Nikpay M, Goel A, Won HH, et al. A comprehensive 1,000 Genomes-based genome-wide association meta-analysis of coronary artery disease. Nat Genet. 2015;47(10):1121–1130.

9. Mars N, Koskela J, Ripatti P, et al. Polygenic and clinical risk scores and their impact on age at onset and prediction of cardiometabolic diseases and common cancers. Nat Med. 2020;26(4).549–557.

10. Koyama S, Ito K, Terao C, et al. Population-specific and trans-ancestry genome-wide analyses identify distinct and shared genetic risk loci for coronary artery disease. Nat Genet. 2020;52(11):1169–1177.

11. Knowles J, Ashley E. Cardiovascular disease: The rise of the genetic risk score. PLOS Medicine. 2018;15(3):e1002546.

12. Torkamani A, Wineinger N, Topol E. The personal and clinical utility of polygenic risk scores. Nature Review Genetics. 2018;19(9):581–590.

13. Wise A, Manolio T, Mensah G, et al. Genomic medicine for undiagnosed diseases. Lancet. 2019;394(10197):533–540.

14. Claussnitzer M, Cho J, Collins R, et al. A brief history of human disease genetics. Nature. 2020;577(7789):179–189.

15. Inoyue M, Abraham G, Nelson C, et al. Genomic Risk Prediction of Coronary Artery Disease in 480,000 Adults: Implications for Primary Prevention. J Am Coll Cardiol. 2018;72(16):1883–1893.

16. Khera AV, Chaffin M, Aragam KG, et al. Genome-wide polygenic scores for common diseases identify individuals with risk equivalent to monogenic mutations. Nat Genet. 2018;50(9):1219–1224.

17. Weale M, Riveros-McKay F, Selzam S, et al. Validation of an Integrated Risk Tool, Including Polygenic Risk Score, for Atherosclerotic Cardiovascualr Disease in Multiple Ethnicities and Ancestries. Am J Cardiol. 2021;148:157–164.

18. Elliott J, Bodinier B, Bond T, et al. Predictive Accuracy of a Polygenic Risk Score-Enhanced Prediction Model vs a Clinical Risk Score for Coronary Artery Disease. JAMA. 2020;323(7):636–645.

19. Mosley J, Gupta D, Tan J, et al. Predictive Accuracy of a Polygenic Risk Score Compared with a Clini-cal Risk Score for Incident Coronary Heart Disease. JAMA. 2020;323(7):627–635.

20. Vilhjálmsson B, Yang J, Finucane H, et al. Modeling Linkage Disequilibrium Increases Accuracy of Polygenic Risk Scores. American Journal of Human Genetics. 2015;97(4):576–592.

21. Ge T, Chen C, Ni Y, et al. Polygenic prediction via Bayesian regression and continuous shrinkage priors. Nature Communications. 2019;10(1).

22. Lloyd-Jones L, Zeng J, Sidorenko J, et al. Improved polygenic prediction by Bayesian multiple regression on summary statistics. Nature Communications. 2019;10(1).

23. Márquez-Luna C, Gazal S, Loh P, et al. LDpred-funct: incorporating functional priors improves polygenic prediction accuracy in UK Biobank and 23andMe data sets. bioRxiv. 2018;375337.

24. Yang S, Zhou X. Accurate and Scalable Construction of Polygenic Scores in Large Biobank Data Sets. American Journal of Human Genetics. 2020;106(5):679–693.

25. Nagai A, Hirata M, Kamatani Y, et al. Overview of the BioBank Japan Project: Study design and pro-file. Journal of Epidemiology. 2017;27(3):S2–S8.

26. Sudlow C, Gallacher J, Allen N, et al. UK biobank: an open access resource for identifying the causes of a wide range of complex diseases of middle and old age. PLoS Med. 2015;12(3):e1001779.

27. UK Biobank. Biomarker assay quality procedures: approaches used to minimize systematic and random errors (and the wider epidemiological implications): version 1.2. https://biobank.ctsu.ox.ac.uk/crys-tal/cyrstal/docs/biomarker_issues.pdf. Published April 2, 2019. Accessed August 10, 2021.

28. Bycroft C, Freeman C, Petkova D, et al. The UK Biobank resource with deep phenotyping and genomic data. Nature. 2108;562(7726):203–209.

29. Wang Y, Guo J, Ni G, et al. Theoretical and empirical quantification of the accuracy of polygenic scores in ancestry divergent populations. Nature Communications. 2020;11:3865.

30. Backman J, Li A, Marcketta A, et al. Exome sequencing and analysis of 454,787 UK Biobank participants. Nature. 2021;599:628–634.

31. Hippisley-Cox J, Coupland C, Vinogradova Y, et al. Predicting cardiovascular risk in England and Wales: prospective derivation and validation of QRISK2. BMJ. 2008;336(7659):1475–1482.

32. Hippisley-Cox J, Coupland C, Brindle P. Development and validation of QRISK3 risk prediction algorithms to estimate future risk of cardiovascular disease: prospective cohort study. BMJ. 2017;357:j2099.

33. UK Biobank. Genotype imputation and genetic association studies of UK Biobank: interim data release. http://www.ukbiobank.ac.uk/wp-content/uploads/2014/04/imputation_documentation_May2015.pdf. Published May 2015. Accessed January 10, 2019.

34. Willer C, Li Y, Abecasis G. METAL: Fast and efficient meta-analysis of genome wide association scans. Bioinformatics. 2010;26(17):2190–2191.

35. Choi S, Mak T, O’Reilly P. Tutorial: a guide to performing polygenic risk score analyses. Nature Protocols. 2020;15(9):2759–2772.

36. Mak T, Porsch R, Choi S, et al. Polygenic scores via penalized regression on summary statistics. Genetic Epidemiology. 2017;41(6):469–480.

37. Wu C, Zhu J, King A, et al. Novel strategy for disease risk prediction incorporating predicted gene expression and DNA methylation data: a multi-phased study of prostate cancer. Cancer Communications. 2021;41(12):1387–1397.

38. SOMERSD. Stata module to calculate Kendall’s tau-a, Somers’ D. and median differences [computer program]. Version S336401: Boston College Department of Economics; 1998.

39. Harrell FE Jr, Califf RM, Pryor DB, Lee KL, Rosati RA. Evaluation the yield of medical tests. JAMA. 1982;247(18):2543–2546.

40. Newson R. Parameters behind “nonparametric” statistics: Kendall’s tau, Somers’ D and median differences. Stata J. 2002;2(1):45–64.

41. Demler O, Paynter N, Cook N. Tests of calibration and goodness-of-fit in the survival setting. Statistics in Medicine. 2015;34(10):1659–1980.

42. Leening M, Vedder M, Witteman J, et al. Net Reclassification Improvement: Computation, Interpretation, and Controversies A Literature Review and Clinician’s Guide. Ann Intern Med. 2014;160:122–131.

43. Pencina MJ, Steyerberg EW, D’Agostino RB Sr. Net reclassification index at event rate: properties and relationships. Stat Med. 2017;36(28):4455–4467.

44. The R Project for Statistical Computing [computer Program]. Version 4.0.0, Vienna, Austria: 2013.

45. Anaconda Software Distribution [Internet]. Anaconda Documentation. Anaconda Inc.; 2020. Available from: https://docs.anaconda.com/

46. Aragam K, Dobbyn A, Judy R. et al. Limitations of Contemporary Guidelines for Managing Patients at High Genetic Risk of Coronary Artery Disease. J Am Coll Cardiol. 2020;75(22):2769–2780.

47. Riveros-McKay F, Weale M, Moore R, et al. Integrated Polygenic Tool Substantially Enhances Coronary Artery Disease Prediction. Circulation: Genomic and Precision Medicine. 2021; 14(2):e003304.

48. Gola D, Erdmann J, Läll K, et al. Population Bias in Polygenic Risk Prediction Models for Coronary Artery Disease. Circulation: Genomic and Precision Medicine. 2020;13(6):e002932.

49. Matsunaga H, Ito K, Akiyama M, et al. Transethnic Meta-Analysis of Genome-Wide Association Studies Identifies Three New Loci and Characterizes Population-Specific Differences for Coronary Artery Disease. Circulation: Genomic and Precision Medicine. 2020;13(3):e002670.

50. Manikpurage H, Eslami A, Perrot N, et al. Polygenic Risk Score for Coronary Artery Disease Improves the Prediction of Early-Onset Myocardial Infarction and Mortality in Men. Circulation: Genomic and Precision Medicine. 2021;14(6):e003452.

51. Thériault S, Lali R, Chong M, et al. Polygenic Contribution in Individuals With Early-Onset Coronary Artery Disease. Circulation: Genomic and Precision Medicine. 2018;11(1).

52. Fry A, Littlejohns T, Sudlow C, et al. Comparison of Sociodemographic and Health-Related Characteristics of UK Biobank Participants with Those fo the General Population. American Journal of Epidemiology. 2017;186(9):1026–1034.

53. Fritsche L, Ma Y, Zhang D, et al. On cross-ancestry cancer polygenic risk scores. PLoS Genet. 2021;17(9):e1009670.

54. Chen C, Han J, Hunter D, Kraft P, Price A. Explicit Modeling of Ancestry Improves Polygenic Risk Scores and BLUP Prediciton. Genetic Epidemiology. 2015;39(6):427–438.

55. Cai M, Xiao J, Zhang S, et al. A unified framework for cross-population trait prediction by leveraging the genetic correlation of polygenic traits. AJHG. 2021;108(4):632–655.

56. Hu YJ, Schmidt AF, Dudbridge F, et al; The GENIUS-CHD Consortium. Impact of selection bias on estimation of subsequent event risk. Circ Cardiovasc Genet. 2017;10(5):e001616.

