## Supplementary for "Polygenic Risk Score Improves the Accuracy of a Clinical Risk Score for Coronary Artery Disease"

### Supplementary Content

**Methods**

**References**

**Table 1.** Definition of variables in UK Biobank

**Table 2.** Definition of Coronary Artery Disease (CAD)

**Table 3.** Descriptive Characteristics

**Table 4.** Association of Different Polygenic Risk Scores (PRS) With Coronary Artery Disease (CAD) in Tuning Case-Control and Prospective Cohort Study

**Table 5.** C Statistics (Derived for Cox Regression) for CAD Using Recalibrated Models in the PCE Prospective Cohort.

**Table 6.** Risk Reclassification at 7.5% Threshold for CAD Using Recalibrated PCE and PCE + PRS Models Stratified by Gender and Age Group (below or above 55 years of age)

**Table 7.** Net Reclassification Improvement and Integrated Discrimination Improvement Results in Secondary Analysis of 6,753 African Ancestry Participants

**Figure 1.** Correlation of PRSs Constructed by Different Methods in the Tuning Set of 9,499 Prevalent CAD cases and an Equal Number of Controls

**Figure 2.** Calibration and Recalibration Plots Polygenic Risk Score for Coronary Artery Disease (CAD) and PRS Combined with PCE, Using a UK Biobank Prospective Cohort Sample

#### Methods

##### Variables included in PCE

Smoking status was based on self-reported information on current smoking status ("most or all days" and "occasional" smokers), previous smoking status and non-smoking status (previous and non-smoking are coded as non-smokers in PCE). Smoking status was determined to be missing when participant's answer to the nurse administered questionnaires was "Prefer not to answer." Mean systolic blood pressure was calculated from two measurements taken seated using an appropriate cuff and an Omron HEM-7015IT digital BP monitor or a manual reading.[3] In cases where digital BP measurement was not available, the manual measurement was utilized. We defined medication status based on nurse administered questionnaires on blood pressure lowering and lipid lowering medications. Serum data was corrected for laboratory dilution effects and excluded if they fell outside the defined UK Biobank specifications.[4] Total serum cholesterol and HDL cholesterol were obtained via enzymatic assays.[4] We calculated their ratio and corrected total and HDL cholesterol for individuals on lipid-lowering medication by dividing total cholesterol by 0.73 and HDL cholesterol by 1.03.[5]

Information on prevalent disease was obtained from nurse administered questionnaires as well as from relevant International Classification of Disease (ICD) or Office of Population Censuses and Surveys Classification of Surgical Operations and Procedures (OPCS-4) codes. These were obtained from in hospital episode statistics (HES) data with a date of event preceding the date of assessment center attendance. Type 1 and 2 diabetes were defined by self-reported diagnoses or from ICD codes in HES data prior to assessment date. In addition, type 2 diabetes was defined by self-reported medications and HbA1c measurements at baseline ≥ 48 mmol/mol. All variables are defined based on ICD-9, ICD-10, OPCS-4, and relevant UK Biobank field codes as shown in Table 1 of the Supplementary.

##### Calculation of Polygenic Risk Scores (PRS)

A PRS is calculated as the weighted sum of an individual's risk alleles. Several factors need to be considered during PRS calculations including (i) the weights used for each SNP, (ii) the number of SNPs included in each score, and (iii) the method to account for correlations between SNPs (linkage disequilibrium [LD]).

We looked at seven methods for PRS calculation that approach these concerns differently:

1. Clumping and Thresholding[6] (P +T) – This method is commonly used and estimates SNP effects from the largest GWAS as the SNP weights. LD is accounted for by a clumping algorithm, which produces a subset of independent SNPs while selecting those weight the strongest association to the desired phenotype. The number of SNPs in the subset included in the PRS is selected by applying a P-value threshold to the original GWAS dataset. Typically, calculations are done with multiple P-value thresholds, and in some cases multiple clumping parameters, to maximize the ability of the PRS to predict the desired target phenotype. Polygenic risk scores were calculated over a range of $r^2$ thresholds (0.2, 0.4, 0.6, 0.8) and P values (1, 0.5, 0.05, 5e-4, 5e-6, 5e-8).

2. LDpred[7] – The LDpred algorithm uses a Bayesian approach to generate posterior mean effects for each SNP included in the GWAS by conditioning on a genetic prior of effects and LD information from an external reference panel. 1000 Genome Project Phase 3 LD reference panels for each population were used. Only the HapMap3 subset of SNPs were used in the tuning of this method. The prior includes two parameters that need to be explored: the heritability explained by the genotype, which is estimated from the GWAS and accounts for sampling noise and LD, and the fraction of causal markers (the fraction of genetic markers with non-zero effects). A LD radius is also required, indicating the number of SNPs adjusted for on each side of a given SNP. In this study, the LD radius was set to 267 and the fraction of causal markers were examined at p = (1, 0.1, 0.3, 1e-2, 3e-2, 1e-3, 3e-3). Like P+T, multiple fractions of causal markers are investigated to optimize the predictive accuracy of the PRS. LD is approximated using an external reference panel that is specific to ancestry of the GWAS dataset.

3. Lassosum[8] - Lassosum aims to achieve maximum predictive accuracy in similar fashion to LDpred, by optimizing SNP weights and accounting for LD. Lassosum performs this by using a penalized regression model to carry out shrinkage and the selection of SNPs from the GWAS dataset, while using the same external LD reference panel as LDpred. Two model parameters (s and lambda) are set, with the PRS being calculated over a grid of s and lambda values in order to maximize phenotypic prediction. For CAD, lassosum has been shown to perform similar to or better than LDpred.[7,8] LD reference panels for each GWAS dataset were defined previously and used as recommended by the author.[9] Reference panels were also set to the hg19 genome.

4. PRS-CS[10] - This method is another method that utilizes a high-dimensional Bayesian framework to infer posterior effect sizes of SNPs using genetic prior effect sizes and an external LD reference panel. In contrast to other methods developed, PRS-CS places a continuous shrinkage (CS) prior on SNP effect sizes. This allows for marker-specific adaptive shrinkage (shrinkage on each genetic marker is adaptive to the strength of the association signal in GWAS) and is more robust to different genetic architectures. PRS-CS is also more accurate in modeling local LD patterns. In simulation, PRS-CS has been shown to outperform LDpred, especially as training sample size increases.[10] For CAD, PRS-CS has demonstrated similar predictive accuracy to LDpred. For our study, the European (EUR) and East Asian (EAS) superpopulation LD reference panels constructed using 1000 Genome Project Phase 3 samples provided by the author were implemented. The European meta-analysis dataset used the EUR LD reference panel, and the Biobank Japan dataset used the EAS reference panel. The polygenic risk scores were calculated using four levels of global shrinkage parameter phi = (1e-6, 1e-4, 1e-2, 1) in order to find the optimal phi value during validation.

5. sBayesR[11] – Another method that infers posterior mean effect sizes from a GWAS dataset and a LD matrix. This method assumes a finite number of normal distributions to account for sparsity with probabilities ranging from 1 to C, the number of components in the mixture model. The recommended parameters for this method are to set C = 4 with parameters $\gamma_c$ = (0,0.01,0.1,1), indicating the scaling factor for the mixture component variance. C was set to 0.95, 0.02, 0.02, and 0.01 in our analysis. This method also requires a shrunk LD matrix, which has been provided by the author. Two different matrices were made available: a 1.1M SNP set constructed by restricting 1,365,446 SNPs from HapMap3 to MAF > 0.01 and a 2.8M SNP set constructed by applying LD-pruning to a larger 8 million variant set from the UK Biobank to MAF > 0.01.

6. LDpred-funct[12] - This method is a modified version of LDpred-funct-inf, which itself is a modification of LDpred-inf. LDpred-funct-inf extends LDpred-inf to incorporate functionally informed priors on causal effect sizes using the baseline-LD model.[13] The 1000 Genome Project Phase 3 derived baseline-LD models (v1.1) containing 75 functional annotations estimated by stratified LD score regression was used for both the European meta-analysis and Japan Biobank datasets. The corresponding EUR and EAS frequency files and HapMap3 subset derived weights were likewise used in the tuning of this method. Each trait is used to determine the per-SNP heritability under the baseline-LD model. LDpred-funct further modifies this method by using cross-validation to regularize posterior mean causal effect sizes. SNPs are ranked by their absolute posterior effect sizes and partitioned into K bins. The relative weights of each bin are determined based on their predictive value in the validation dataset. This non-parametric shrinkage allows LDpred-funct to optimize predictive accuracy regardless of the underlying genetic architecture.

7. DBSLMM[14] – Deterministic Bayesian Sparse Linear Mixed Model (DBSLMM) is a method designed to create a scalable PRS method that is computationally less intensive and thus can be used for larger GWAS datasets efficiently. This method relies on the same effect size distribution assumption made in the BSLMM method. A major difference between these two methods is the use of Markov chain Monte Carlo (MCMC) for posterior sampling in BSLMM, leading it to be computationally slow especially as GWAS size increases. DBSLMM instead utilizes a searching algorithm to subset SNPs with potential large effect sizes. Small effect size SNPs are combined to find total effect size. Both subsets are then combined with a block-diagonal SNP correlation matrix. This leads to DBSLMM having a computationally complexity approximately linear respective to both sample size and the number of SNPs. We tuned this method using the EUR and EAS region reference LD panels in a similar manner as other methods. The recommended block information used in the

derivation of the polygenic risk scores was obtained from a previously published study.[7] We tested DBSLMM at three folds of heritability levels: 0.8, 1.0. and 1.2, respectively.

**Reclassification Metrics**

We calculated the net reclassification index (NRI) at the current recommended threshold for treatment in the US (7.5%). The associated category-free NRI and integrated discrimination improvement (IDI) were also calculated. The NRI is calculated for nested models, PCE vs PCE + PRS in our study, using a 2x2 classification table at the defined threshold. "Upward" movement in categories is defined as subjects with a better outcome after reclassification while "downward" movement is defined as subjects with a worse reclassification. The improvement in reclassification is calculated as the sum of the differences in proportions in subjects moving up minus those moving down that had an event/outcome, and the proportion of subjects moving down minus those moving up that did not experience the outcome.[15] NRI has been shown to be sensitive to the number and choices of thresholds[16], and as such the category-free (continuous) NRI has been proposed. The continuous NRI is calculated as the relative increase in the predictive probabilities of subjects that experience the event and the decrease in those who do not.[17] IDI is defined as a measure that integrates the NRI over all possible cut-offs for the probability of the outcome. The IDI is equivalent to the difference in the discrimination slopes between 2 models.[15]

#### References


1. Wang Y, Guo J, Ni G, et al. Theoretical and empirical quantification of the accuracy of polygenic scores in ancestry divergent populations. *Nature Communications*. 2020; 11:3865.
2. Backman J, Li A, Marcketta A, et al. Exome sequencing and analysis of 454,787 UK Biobank participants. *Nature*. 2021; 599:628-634.
3. UK Biobank Coordinating Centre; UK Biobank: Protocol for a large-scale prospective epidemiological resource. 2007; http://www.ukbiobank.ac.uk/wp-content/uploads/2011/11/UK-Biobank-Protocol.pdf.
4. UK Biobank. Biomarker assay quality procedures: approaches used to minimize systematic and random errors (and the wider epidemiological implications). 2019; https://biobank.ctsu.ox.ac.uk/showcase/docs/biomarker_issues.pdf.
5. Nissen SE, Tuzcu EM, Schoenhagen P, et al. Statin therapy, LDL cholesterol, C-reactive protein, and coronary artery disease. N Engl J Med. 2005; 352(1):29-38.
6. Choi SW, Mak TS, O'Reily PF. Tutorial: a guide to performing polygenic risk score analyses. Nature Protocols. 2020; 15:2759-2772.
7. Vilhjalmsson BJ, Yang J, Finucane HK, et al. Modeling Linkage Disequilibrium Increase Accuracy of Polygenic Risk Scores. Am J Hum Genet. 2015; 97(4):576-592.
8. Mak TSH, Porsch RM, Choi SW, Zhou X, Sham PC. Polygenic scores via penalized regression on summary statistics. Genetic epidemiology. 2017; 41(6):469-480.
9. Berisa T and Pickrell J. Approximately independent linkage disequilibrium blocks in human populations. Bioinformatics. 2016, 32(2):283-285.
10. Ge T, Chen CY, Fang YA, Smoller JW. Polygenic prediction via Bayesian regression and continuous shrinkage priors. Nature Communications. 2019; 10(1):1776.
11. Lloyd-Jones LR, Zeng J, Sidorenko J, et al. Improved polygenic prediction by Bayesian multiple regression on summary statistics. Nature Commuinications. 2019; 10:5086.
12. Márquez-Luna C, Gazal S, Loh P, et al. LDpred-funct: incorporating functional priors improves polygenic prediction accuracy in UK Biobank and 23andMe data sets. BioRxiv. 2018; doi: https://doi.org/10.1101/375337.
13. Gazal S, Finucane H, Furlotte A, et al. Linkage disequilibrium-dependent architecture of human complex traits shows action of negative selection. Nature Genetics. 2017; 49:1421 EP-,09.
14. Sheng Y, Zhou X. Accurate and Scalable Construction of Polygenic Scores in Large Biobank Data Sets. Am J Hum Genet. 2020; 106(5):679-693.
15. Goodard M, et al. In: Balding D, Moltke I, Marioni J, editors. Handbook of Statistical Genomics. 4 ed. Hoboken, USA: John Wiley and Sons; 2019; p. 799-20.
16. Steyerberg EW, Vickers AJ, Cook NR, et al. Assessing the performance of prediction models: a framework for traditional and novel measures. Epidemiology. 2010; 21(1):128-138.
17. Tzoulaki I, Liberopoulos G, Ioannidis JP. Use of reclassification for assessment of improved prediction: an empirical evaluation. International journal of epidemiology. 2011; 40(4):1094-1105.

**Table 1.** Definition of Variables in UK Biobank

| | HES statistics ICD-10 and ICD-9 codes (with date of event < date of visit) | Self-reported Data-field 20002 | Treatment and medication code Data-Field 20003 | Other data fields |
|---|---|---|---|---|
| **Type 1 diabetes** | ICD-10 E10, O230; ICD-9 25001, 25011, 25021, 25031, 25041, 25051, 25061, 25071, 25081, 25091, 25003, 25013, 25023, 25033, 25043, 25053, 25063, 25073, 25083, 25093 | 1222 | | |
| **Type 2 diabetes** | ICD-10 E11, O231; ICD-9 25000, 25010, 25020, 25030, 25040, 25050, 25060, 25070, 25080, 25090, 25002, 25012, 25022, 25032, 25042, 25052, 25062, 25072, 25082, 25092 | 1223, 1220 | 1140868902, 1140874646, 1140874674, 1140874718, 1140874744, 1140883066, 1140884600, 1141152590, 1141157284, 1141168660, 1141171646, 1141173882, 1141189090 | Biobank field 2443 = 1; HbA1c measurement ≥ 48 mmol/mol |
| **Lipid lowering medication** | | | 1140861954, 1140861958, 1140888594, 1140888648, 1141146234, 1141192410, 1141192736 | |
| **Blood pressure lowering medication** | | | 1140860192, 1140860292, 1140860696, 1140860728, 1140860750, 1140860806, 1140860882, 1140860904, 1140861088, 1140861190, 1140861276, 1140866072, 1140866078, 1140866090, 1140866102, 1140866108, 1140866122, 1140866138, 1140866156, 1140866162, 1140866724, 1140866738, 1140868618, 1140872568, 1140874706, 1140874744, 1140875808, 1140879758, 1140879760, 1140879762, 1140879802, 1140879806, 1140879810, 1140879818, 1140879822, 1140879826, 1140879830, 1140879834, 1140879842, 1140879866, 1140884298, 1140888552, 1140888556, 1140888560, 1140888646, 1140909706, 1140910442, 1140910614, 1140916356, 1140923272, 1140923336, 1140923404, 114923712, 1140926778, 1140928226, 1141145660, 1141146126, 1141152998, 1141153026, 1141164276, 1141165470, 1141166006, 1141169516, 1141171336, 1141180592, 1141180772, 1141180778, 1141184722, 1141193282, 1141194794, 1141194810 | Biobank fields 6177, 6153 = 2 |

**Table 2.** Definition of Coronary Artery Disease (CAD)

| **ICD-10** | **ICD-9** | **OPCS-4** | **Non-cancer illness code (Biobank field: 20002)** | **Operation code (Biobank field: 20004)** | **Vascular/heart problems diagnosed by doctor (Biobank field: 6150)** |
|---|---|---|---|---|---|
| I21 | 410 | K40.1-4 | 1075: Heart attack/myocardial infarction | 1070: Coronary angioplasty | 1: Heart attack |
| I22 | 411 | K41.1-4 | | 1095: Coronary artery bypass graft | |
| I23 | 412 | K45.1-5 | | | |
| I24.1 | | K49.1-2 | | | |
| I25.2 | | K49.8-9 | | | |
| | | K50.2 | | | |
| | | K75.1-4 | | | |
| | | K75.8-9 | | | |

**Table 3A.** Descriptive Characteristics of Tuning (Case-Control) Set (N=18,998)

| | Men | Women |
|---|---|---|
| **N** | 12043 | 6955 |
| **Age (years) (mean (SD)) *** | 60.23 (7.21) | 58.16 (7.82) |
| **Mean SBP (mmHg) (mean (SD)) *** | 137.92 (18.57) | 137.62 (18.7) |
| **Smoking category (%) *** | | |
| **Non-smoker** | 4652 (38.6) | 3857 (55.5) |
| **Ex-smoker** | 5974 (49.6) | 2450 (35.2) |
| **Current smoker** | 1417 (11.8) | 648 (9.3) |
| **Type 1 diabetes (%) *** | 325 (2.7) | 118 (1.7) |
| **Type 2 diabetes (%) *** | 2621 (21.8) | 775 (11.1) |
| **Blood pressure lowering medication (%) *** | 7877 (65.4) | 2503 (36.0) |
| **Cholesterol (mmol/L) (mean (SD)) *** | 4.82 (1.15) | 5.63 (1.18) |
| **HDL cholesterol (mmol/L) (mean (SD)) *** | 1.2 (0.3) | 1.55 (0.38) |
| **Lipid lowering medication (%) *** | 7230 (60.0) | 1886 (27.1) |

* Variables used in PCE. Current smokers vs non-current smokers and diabetes as a binary variable

**Table 3B.** Descriptive Characteristics of PCE Prospective Cohort/Testing Set (N=272,307)

| | Full Population | | Incident CAD | |
|---|---|---|---|---|
| | **Men** | **Women** | **Men** | **Women** |
| **N** | 124118 | 148189 | 5093 | 1943 |
| **Age (years) (Mean (SD))** | 56.8 (8.09) | 56.6 (7.88) | 60.44 (6.6) | 61.1 (6.46) |
| **Mean SBP (mmHg) (mean (SD))** | 137.86 (18.66) | 137.84 (18.68) | 138.32 (18.74) | 138.13 (18.45) |
| **Smoking category (%)** | | | | |
| **Current** | 14580 (11.7) | 12747 (8.6) | 806 (15.8) | 325 (16.7) |
| **Non-current** | 109538 (88.3) | 135442 (91.4) | 4287 (84.2) | 1618 (83.3) |
| **Type 1 Diabetes (%)** | 1260 (1.0) | 1016 (0.7) | 150 (2.9) | 59 (3.0) |
| **Type 2 Diabetes (%)** | 12832 (10.3) | 8962 (6.0) | 1082 (21.2) | 345 (17.8) |
| **Blood pressure lowering medication (%)** | 21506 (17.3) | 15629 (10.5) | 2220 (43.6) | 814 (41.9) |
| **Cholesterol (mmol/L) (mean (SD))** | 5.57 (1.1) | 5.91 (1.12) | 5.28 (1.58) | 5.71 (1.62) |
| **HDL cholesterol (mmol/L) (mean (SD))** | 1.29 (0.31) | 1.6 (0.38) | 1.23 (0.29) | 1.49 (0.36) |
| **Person-years of observation (mean (SD))** | 11.79 (2.27) | 12.13 (1.65) | 4.82 (2.51) | 5.15 (2.5) |
| **Lipid lowering medication (%)** | 21562 (17.4) | 15611 (10.5) | 1466 (28.8) | 479 (24.7) |

**Table 3C.** Descriptive Characteristics of the PCE Prospective Cohort and Excluded Participants

| | **Prospective cohort (N = 272307)** | | **Excluded (N = 43474) *** | | |
|---|---|---|---|---|---|
| | Men | Women | Men | Women | Missing counts |
| **N** | 124118 | 148189 | 17790 | 20084 | 5600 |
| **Age (years) (Mean (SD))** | 56.8 (8.09) | 56.6 (7.88) | 57.03 (8.08) | 56.65 (7.89) | |
| **Mean SBP (mmHg) (mean (SD))** | 137.86 (18.66) | 137.84 (18.68) | 137.84 (18.72) | 137.85 (7.89) | 633 |
| **Smoking category (%)** | | | | | 135 |
| **Current** | 14580 (11.7) | 12747 (8.6) | 2088 (11.8) | 1718 (8.5) | |
| **Non-current** | 109538 (88.3) | 135442 (91.4) | 15621 (88.2) | 18366 (91.5) | |
| **Type 1 Diabetes (%)** | 1260 (1.0) | 1016 (0.7) | 211 (1.2) | 161 (0.8) | |
| **Type 2 Diabetes (%)** | 12832 (10.3) | 8962 (6.0) | 2026 (11.4) | 1257 (6.3) | |
| **Blood pressure lowering medication (%)** | 21506 (17.3) | 15629 (10.5) | 5159 (29.1) | 4077 (20.3) | |
| **Cholesterol (mmol/L) (mean (SD))** | 5.57 (1.1) | 5.91 (1.12) | 5.5 (1.13) | 5.89 (1.13) | 15553 |
| **HDL cholesterol (mmol/L) (mean (SD))** | 1.29 (0.31) | 1.6 (0.38) | 1.28 (0.31) | 1.6 (0.38) | 42489 |
| **Person-years of observation (mean (SD))** | 11.79 (2.27) | 12.13 (1.65) | 10.39 (6.23) | 11.39 (4.53) | |
| **Lipid lowering medication (%)** | 21562 (17.4) | 15611 (10.5) | 3776 (21.3) | 2285 (11.4) | |

* 5600 participants excluded from calculations due to missing variables on gender

**Table 3D.** Descriptive Characteristics of Tuning (Case-Control) Set, Secondary Analysis (N=218)

| | **Men** | **Women** |
|---|---|---|
| **N** | 111 | 107 |
| **Age (years) (mean (SD)) *** | 55.3 (8.46) | 52.92 (8.17) |
| **Mean SBP (mmHg) (mean (SD)) *** | 135.09 (18.69) | 138.38 (19.36) |
| **Smoking category (%) *** | | |
| **Non-smoker** | 56 (50.5) | 81 (75.7) |
| **Ex-smoker** | 32 (28.8) | 13 (12.15) |
| **Current smoker** | 23 (20.7) | 13 (12.15) |
| **Type 1 diabetes (%) *** | 2 (1.8) | 5 (4.7) |
| **Type 2 diabetes (%) *** | 48 (43.2) | 38 (35.5) |
| **Blood pressure lowering medication (%) *** | 70 (63.1) | 54 (50.5) |
| **Cholesterol (mmol/L) (mean (SD)) *** | 4.29 (1.47) | 4.56 (1.25) |
| **HDL cholesterol (mmol/L) (mean (SD)) *** | 1.25 (0.28) | 1.5 (0.33) |
| **Lipid lowering medication (%) *** | 49 (44.1) | 36 (33.6) |

* Variables used in PCE. Current smokers vs non-current smokers and diabetes as a binary variable

**Table 3E.** Descriptive Characteristics of PCE Prospective Cohort/Testing Set, Secondary Analysis (N= 6,753)

| | Full Population | | Incident CAD | |
|---|---|---|---|---|
| | **Men** | **Women** | **Men** | **Women** |
| **N** | 2901 | 3852 | 46 | 42 |
| **Age (years) (Mean (SD))** | 51.26 (8.13) | 51.9 (7.93) | 55.61 (8.57) | 55.4 (8.41) |
| **Mean SBP (mmHg) (mean (SD))** | 136. 87 (18.56) | 138.12 (18.59) | 136.83 (16.83) | 135.08 (20.25) |
| **Smoking category (%)** | | | | |
| **Current** | 481 (16.6) | 356 (9.2) | 10 (21.7) | 6 (14.3) |
| **Non-current** | 2420 (83.4) | 3496 (90.8) | 36 (78.3) | 36 (85.7) |
| **Type 1 Diabetes (%)** | 57 (2) | 67 (1.7) | 7 (15.2) | 3 (7.1) |
| **Type 2 Diabetes (%)** | 606 (20.1) | 699 (18.1) | 21 (45.7) | 26 (62.9) |
| **Blood pressure lowering medication (%)** | 828 (28.5) | 1325 (34.4) | 23 (50) | 27 (64.3) |
| **Cholesterol (mmol/L) (mean (SD))** | 4.99 (1.26) | 5.17 (1.18) | 4.77 (1.39) | 4.95 (1.53) |
| **HDL cholesterol (mmol/L) (mean (SD))** | 1.3 (0.33) | 1.54 (0.36) | 1.32 (0.37) | 1.38 (0.38) |
| **Person-years of observation (mean (SD))** | 12.69 (1.8) | 12.73 (1.6) | 5.48 (2.21) | 5.22 (2.41) |
| **Lipid lowering medication (%)** | 395 (13.6) | 462 (12) | 13 (28.3) | 12 (28.6) |

**Table 3F.** Descriptive Characteristics of the PCE Prospective Cohort and Excluded Participants, Secondary Analysis

| | Prospective cohort (N = 6753) | | Excluded (N = 1135) * | | |
|---|---|---|---|---|---|
| | Men | Women | Men | Women | Missing counts |
| **N** | 2901 | 3852 | 425 | 552 | 158 |
| **Age (years) (Mean (SD))** | 51.26 (8.13) | 51.9 (7.93) | 51.99 (8.49) | 51.56 (7.97) | |
| **Mean SBP (mmHg) (mean (SD))** | 136. 87 (18.56) | 138.12 (18.59) | 137.14 (18.4) | 137.95 (18.96) | 23 |
| **Smoking category (%)** | | | | | 26 |
| **Current** | 481 (16.6) | 356 (9.2) | 68 (16) | 70 (12.7) | |
| **Non-current** | 2420 (83.4) | 3496 (90.8) | 357 (84) | 482 (87.3) | |
| **Type 1 Diabetes (%)** | 57 (2) | 67 (1.7) | 8 (1.9) | 10 (1.8) | |
| **Type 2 Diabetes (%)** | 606 (20.1) | 699 (18.1) | 106 (24.9) | 104 (18.8) | |
| **Blood pressure lowering medication (%)** | 828 (28.5) | 1325 (34.4) | 152 (35.8) | 177 (32.1) | |
| **Cholesterol (mmol/L) (mean (SD))** | 4.99 (1.26) | 5.17 (1.18) | 4.9 (1.27) | 5.16 (1.15) | 504 |
| **HDL cholesterol (mmol/L) (mean (SD))** | 1.3 (0.33) | 1.54 (0.36) | 1.28 (0.3) | 1.54 (0.36) | 1058 |
| **Person-years of observation (mean (SD))** | 12.69 (1.8) | 12.73 (1.6) | 12.16 (3.4) | 12.69 (2.11) | |
| **Lipid lowering medication (%)** | 395 (13.6) | 462 (12) | 68 (16) | 60 (10.9) | |

* 158 participants excluded from calculations due to missing variables on gender

**Table 4A.** Association of Different Polygenic Risk Scores (PRS) With Coronary Artery Disease (CAD) in Tuning Case-Control Study and in Prospective Cohort Study in European GWAS meta-analysis

| | | AUC (95% CI) PRS | |
|---|---|---|---|
| **Methods** | **P-value/Tuning Parameters** | **Case-control study (N = 9,499)** | **Cohort Study (N = 7,036)** |
| Clumping and Thresholding | R2=0.2, P=0.00000005 | 0.6179 (0.6137-0.6221) | 0.595 (0.5917-0.5982) |
| Clumping and Thresholding | R2=0.2, P=0.000005 | 0.6225 (0.6183-0.6267) | 0.5955 (0.5922-0.5987) |
| Clumping and Thresholding | R2=0.2, P=0.0005 | 0.6230 (0.6188-0.6272) | 0.5976 (0.5944-0.6009) |
| Clumping and Thresholding | R2=0.2, P=0.05 | 0.6005 (0.5963-0.6047) | 0.5835 (0.5802-0.5868) |
| Clumping and Thresholding | R2=0.2, P=0.5 | 0.5966 (0.5924-0.6047) | 0.578 (0.5747-0.5813) |
| Clumping and Thresholding | R2=0.2, P=1 | 0.5962 (0.592-0.6004) | 0.5777 (0.5744-0.5809) |
| Clumping and Thresholding | R2=0.4, P=0.00000005 | 0.6155 (0.6113-0.6197) | 0.593 (0.5897-0.5963) |
| Clumping and Thresholding | R2=0.4, P=0.000005 | 0.6221 (0.6179-.06263) | 0.596 (0.5927-0.5993) |
| Clumping and Thresholding | R2=0.4, P=0.0005 | 0.6291 (0.6249-0.6333) | 0.6039 (0.6007-0.6072) |
| Clumping and Thresholding | R2=0.4, P=0.05 | 0.6146 (0.6104-0.6188) | 0.5941 (0.5908-0.5974) |
| Clumping and Thresholding | R2=0.4. P=0.5 | 0.61 (0.6058-0.6142) | 0.5871 (0.5838-0.5903) |
| Clumping and Thresholding | R2=0.4, P=1 | 0.6094 (0.6052-0.6136) | 0.5865 (0.5832-0.5897) |
| Clumping and Thresholding | R2=0.6, P=0.00000005 | 0.6138 (0.6096-0.6180) | 0.5911 (0.5878-0.5944) |
| Clumping and Thresholding | R2=0.6, P=0.000005 | 0.6236 (0.6194-0.6278) | 0.5982 (0.5949-0.6014) |
| Clumping and Thresholding | R2=0.6, P=0.0005 | 0.6314 (0.6272-0.6356) | 0.6043 (0.6010-0.6075) |
| Clumping and Thresholding | R2=0.6, P=0.05 | 0.6230 (0.6188-0.6272) | 0.5997 (0.5964-0.603) |
| Clumping and Thresholding | R2=0.6, P=0.5 | 0.6169 (0.6127-0.6211) | 0.5924 (0.5892-0.5957) |
| Clumping and Thresholding | R2=0.6, P=1 | 0.6159 (0.6117-0.6201) | 0.5918 (0.5885-0.5951) |
| Clumping and Thresholding | R2=0.8, P=0.00000005 | 0.6069 (0.6027-0.6111) | 0.5852 (0.5819-0.5983) |
| Clumping and Thresholding | R2=0.8, P=0.000005 | 0.6188 (0.6146-0.6230) | 0.5951 (0.5918-0.5983) |
| Clumping and Thresholding | R2=0.8, P=0.0005 | 0.6309 (0.6267-0.6335) | 0.6049 (0.6016-0.6082) |
| Clumping and Thresholding | R2=0.8, P=0.05 | 0.6293 (0.6251-0.6335) | 0.6056 (0.6023-0.6088) |
| Clumping and Thresholding | R2=0.8, P=0.5 | 0.6226 (0.6184-0.6268) | 0.5977 (0.5944-0.6009) |
| Clumping and Thresholding | R2=0.8, P=1 | 0.6217 (0.6175-0.6259) | 0.5972 (0.5939-0.6004) |
| LDpred | p = 0.001 | 0.5322 (0.4874-0.5769) | 0.5198 (0.4806-0.559) |
| LDpred | p = 0.01 | 0.5668 (0.5221-0.6115) | 0.5225 (0.5133-0.5917) |
| LDpred | p = 0.1 | 0.6496 (0.6048-0.6943) | 0.6238 (0.5846-0.663) |

| Methods | P-value/Tuning Parameters | Case-control study (N = 9,499) | Cohort Study (N = 7,036) |
|---|---|---|---|
| LDpred | p = 1 | 0.6356 (0.5909-0.686) | 0.6123 (0.5731-0.6515) |
| LDpred | p = 0.003 | 0.5323 (0.4876-0.577) | 0.5222 (0.483-0.5614) |
| LDpred | p = 0.03 | 0.6585 (0.6138-0.7033) | 0.6316 (0.5924-0.6708) |
| LDpred | p = 0.3 | 0.6413 (0.5966-0.686) | 0.6168 (0.5776-0.6560) |
| lassosum | s=0.2, lambda=0.001 | 0.5826 (0.5737-0.5916) | 0.5681 (0.5602-0.5761) |
| lassosum | s=0.2, lambda=0.0013 | 0.5885 (0.5795-0.5974) | 0.5732 (0.5652-0.5812) |
| lassosum | s=0.2, lambda=0.0016 | 0.5962 (0.5873-0.6052) | 0.5818 (0.5738-0.5898) |
| lassosum | s=0.2, lambda=0.0021 | 0.6065 (0.5976-0.6155) | 0.5905 (0.5825-0.5985) |
| lassosum | s=0.2, lambda=0.0026 | 0.6201 (0.6111-0.629) | 0.5997 (0.5917-0.6077) |
| lassosum | s=0.2, lambda=0.0034 | 0.6346 (0.6257-0.6436) | 0.6133 (0.6053-0.6212) |
| lassosum | s=0.2, lambda=0.0043 | 0.6487 (0.6397-0.6576) | 0.6225 (0.6146-0.6305) |
| lassosum | s=0.2, lambda=0.0055 | 0.6458 (0.6369-0.6548) | 0.6176 (0.6097-0.6256) |
| lassosum | s=0.2, lambda=0.007 | 0.6281 (0.6191-0.637) | 0.6043 (0.5963-0.6122) |
| lassosum | s=0.2, lambda=0.0089 | 0.6106 (0.6017-0.6196) | 0.5903 (0.5824-0.5983) |
| lassosum | s=0.2, lambda=0.0113 | 0.5955 (0.5865-0.6045) | 0.5778 (0.5698-0.5857) |
| lassosum | s=0.2, lambda=0.0114 | 0.5819 (0.573-0.5909) | 0.5648 (0.5568-0.5728) |
| lassosum | s=0.2, lambda=0.0183 | 0.5714 (0.5624-0.5804) | 0.5545 (0.5465-0.5624) |
| lassosum | s=0.2, lambda=0.0234 | 0.5655 (0.5566-0.5745) | 0.5486 (0.5406-0.5565) |
| lassosum | s=0.2, lambda=0.0298 | 0.5577 (0.5488-0.5667) | 0.5411 (0.5332-0.5491) |
| lassosum | s=0.2, lambda=0.0379 | 0.5569 (0.548-0.5659) | 0.541 (0.533-0.549) |
| lassosum | s=0.2, lambda=0.0483 | 0.5311 (0.5221-0.54) | 0.5191 (0.5111-0.527) |
| lassosum | s=0.2, lambda=0.0616 | 0.5311 (0.5221-0.54) | 0.5191 (0.5111-0.527) |
| lassosum | s=0.2, lambda=0.0785 | 0.5311 (0.5221-0.54) | 0.5191 (0.5111-0.527) |
| lassosum | s=0.2, lambda=0.1 | 0.5311 (0.5221-0.54) | 0.5191 (0.5111-0.527) |
| lassosum | s=0.5, lambda=0.001 | 0.6016 (0.5927-0.6106) | 0.5842 (0.5762-0.5921) |
| lassosum | s=0.5, lambda=0.0013 | 0.6069 (0.598-0.6159) | 0.589 (0.581-0.5969) |
| lassosum | s=0.5, lambda=0.0016 | 0.6131 (0.6041-0.6221) | 0.5951 (0.5871-0.6031) |
| lassosum | s=0.5, lambda=0.0021 | 0.6214 (0.6125-0.6304) | 0.6019 (0.594-0.6099) |

| Methods | P-value/Tuning Parameters | Case-control study (N = 9,499) | Cohort Study (N = 7,036) |
|---|---|---|---|
| lassosum | s=0.5, lambda=0.0026 | 0.6328 (0.6239-0.6418) | 0.6106 (0.6026-0.6185) |
| lassosum | s=0.5, lambda=0.0034 | 0.6452 (0.6362-0.6541) | 0.6216 (0.6136-0.6296) |
| lassosum | s=0.5, lambda=0.0043 | 0.6538 (0.6449-0.6628) | 0.626 (0.6181-0.634) |
| lassosum | s=0.5, lambda=0.0055 | 0.6454 (0.6364-0.6543) | 0.6178 (0.6099-0.6258) |
| lassosum | s=0.5, lambda=0.007 | 0.6273 (0.6183-0.6363) | 0.6031 (0.5952-0.6111) |
| lassosum | s=0.5, lambda=0.0089 | 0.6103 (0.6013-0.6193) | 0.5895 (0.5815-0.5974) |
| lassosum | s=0.5, lambda=0.0113 | 0.5946 (0.5856-0.6036) | 0.5767 (0.5688-0.5847) |
| lassosum | s=0.5, lambda=0.0114 | 0.5803 (0.5714-0.5893) | 0.5633 (0.5554-0.5713) |
| lassosum | s=0.5, lambda=0.0183 | 0.5702 (0.5613-0.5792) | 0.5535 (0.5456-0.5615) |
| lassosum | s=0.5, lambda=0.0234 | 0.5645 (0.5556-0.5735) | 0.5476 (0.5397-0.5556) |
| lassosum | s=0.5, lambda=0.0298 | 0.5578 (0.5488-0.5667) | 0.5411 (0.5332-0.5491) |
| lassosum | s=0.5, lambda=0.0379 | 0.5576 (0.5486-0.5665) | 0.5411 (0.5331-0.5491) |
| lassosum | s=0.5, lambda=0.0483 | 0.5311 (0.5221-0.54) | 0.5191 (0.5111-0.527) |
| lassosum | s=0.5, lambda=0.0616 | 0.5311 (0.5221-0.54) | 0.5191 (0.5111-0.527) |
| lassosum | s=0.5, lambda=0.0785 | 0.5311 (0.5221-0.54) | 0.5191 (0.5111-0.527) |
| lassosum | s=0.5, lambda=0.1 | 0.5311 (0.5221-0.54) | 0.5191 (0.5111-0.527) |
| lassosum | s=0.9, lambda=0.001 | 0.6279 (0.619-0.6369) | 0.6056 (0.5977-0.6136) |
| lassosum | s=0.9, lambda=0.0013 | 0.6322 (0.6232-0.6411) | 0.6094 (0.6014-0.6173) |
| lassosum | s=0.9, lambda=0.0016 | 0.6375 (0.6285-0.6464) | 0.6139 (0.606-0.6219) |
| lassosum | s=0.9, lambda=0.0021 | 0.6438 (0.6349-0.6528) | 0.6193 (0.6113-0.6272) |
| lassosum | s=0.9, lambda=0.0026 | 0.6513 (0.6423-0.6602) | 0.6253 (0.6174-0.6333) |
| lassosum | s=0.9, lambda=0.0034 | 0.6563 (0.6474-0.6653) | 0.6293 (0.6214-0.6373) |
| lassosum | s=0.9, lambda=0.0043 | 0.6516 (0.6427-0.6606) | 0.6241 (0.6162-0.6321) |
| lassosum | s=0.9, lambda=0.0055 | 0.6366 (0.6276-0.6455) | 0.611 (0.603-0.6189) |
| lassosum | s=0.9, lambda=0.007 | 0.6193 (0.6103-0.6282) | 0.5958 (0.5879-0.6038) |
| lassosum | s=0.9, lambda=0.0089 | 0.603 (0.5941-0.612) | 0.5823 (0.5743-0.5903) |
| lassosum | s=0.9, lambda=0.0113 | 0.5884 (0.5794-0.5973) | 0.5699 (0.5619-0.5779) |
| lassosum | s=0.9, lambda=0.0114 | 0.576 (0.5671-0.585) | 0.5579 (0.5499-0.5659) |

| Methods | P-value/Tuning Parameters | Case-control study (N = 9,499) | Cohort Study (N = 7,036) |
|---|---|---|---|
| lassosum | s=0.9, lambda=0.0183 | 0.5662 (0.5573-0.5752) | 0.5494 (0.5415-0.5574) |
| lassosum | s=0.9, lambda=0.0234 | 0.5619 (0.553-0.5709) | 0.5448 (0.5368-0.5527) |
| lassosum | s=0.9, lambda=0.0298 | 0.558 (0.549-0.5669) | 0.5412 (0.5332-0.5491) |
| lassosum | s=0.9, lambda=0.0379 | 0.5578 (0.5488-0.5667) | 0.5411 (0.5332-0.5491) |
| lassosum | s=0.9, lambda=0.0483 | 0.5311 (0.5221-0.54) | 0.5191 (0.5111-0.527) |
| lassosum | s=0.9, lambda=0.0616 | 0.5311 (0.5221-0.54) | 0.5191 (0.5111-0.527) |
| lassosum | s=0.9, lambda=0.0785 | 0.5311 (0.5221-0.54) | 0.5191 (0.5111-0.527) |
| lassosum | s=0.9, lambda=0.1 | 0.5311 (0.5221-0.54) | 0.5191 (0.5111-0.527) |
| lassosum | s=1, lambda=0.001 | 0.6221 (0.6132-0.6311) | 0.5982 (0.5902-0.6061) |
| lassosum | s=1, lambda=0.0013 | 0.6231 (0.6141-0.632) | 0.5991 (0.5911-0.607) |
| lassosum | s=1, lambda=0.0016 | 0.6239 (0.6149-0.6329) | 0.5999 (0.5919-0.6078) |
| lassosum | s=1, lambda=0.0021 | 0.6246 (0.6156-0.6335) | 0.6002 (0.5922-0.6082) |
| lassosum | s=1, lambda=0.0026 | 0.6239 (0.6149-0.6328) | 0.5996 (0.5917-0.6076) |
| lassosum | s=1, lambda=0.0034 | 0.6186 (0.6096-0.6276) | 0.5949 (0.587-0.6029) |
| lassosum | s=1, lambda=0.0043 | 0.6086 (0.5996-0.6176) | 0.5864 (0.5785-0.5944) |
| lassosum | s=1, lambda=0.0055 | 0.5972 (0.5882-0.6061) | 0.5776 (0.5697-0.5856) |
| lassosum | s=1, lambda=0.007 | 0.5869 (0.5779-0.5958) | 0.5691 (0.5612-0.5771) |
| lassosum | s=1, lambda=0.0089 | 0.577 (0.5681-0.586) | 0.5598 (0.5519-0.5678) |
| lassosum | s=1, lambda=0.0113 | 0.571 (0.5621-0.58) | 0.5529 (0.545-0.5609) |
| lassosum | s=1, lambda=0.0114 | 0.5662 (0.5572-0.5752) | 0.5471 (0.5392-0.5551) |
| lassosum | s=1, lambda=0.0183 | 0.5606 (0.5516-0.5696) | 0.5427 (0.5348-0.5507) |
| lassosum | s=1, lambda=0.0234 | 0.5593 (0.5503-0.5683) | 0.5418 (0.5339-0.5498) |
| lassosum | s=1, lambda=0.0298 | 0.5584 (0.5494-0.5674) | 0.5412 (0.5332-0.5491) |
| lassosum | s=1, lambda=0.0379 | 0.5578 (0.5488-0.5667) | 0.5411 (0.5332-0.5491) |
| lassosum | s=1, lambda=0.0483 | 0.5311 (0.5221-0.54) | 0.5191 (0.5111-0.527) |
| lassosum | s=1, lambda=0.0616 | 0.5311 (0.5221-0.54) | 0.5191 (0.5111-0.527) |
| lassosum | s=1, lambda=0.0785 | 0.5311 (0.5221-0.54) | 0.5191 (0.5111-0.527) |
| lassosum | s=1, lambda=0.1 | 0.5311 (0.5221-0.54) | 0.5191 (0.5111-0.527) |

| **Methods** | **P-value/Tuning Parameters** | **Case-control study (N = 9,499)** | **Cohort Study (N = 7,036)** |
|---|---|---|---|
| PRS-CS | phi = 0.000001 | 0.6379 (0.6071-0.6688) | 0.6120 (0.5877-0.6363) |
| PRS-CS | phi =0.0001 | 0.6621 (0.6312-0.6929) | 0.6315 (0.6072-0.6558) |
| PRS-CS | phi = 0.01 | 0.6393 (0.6084-0.6701) | 0.6147 (0.5904-0.6390) |
| PRS-CS | phi = 1 | 0.6146 (0.5837-0.6454) | 0.5942 (0.5698-0.6185) |
| sBayesR | LD matrix = HapMap3 and 2.8M | 0.5359 (0.5297-0.5421) | 0.5227(0.5053-0.5400) |
| sBayesR | LD matrix = 2.8M variants | 0.5349 (0.5287-0.5411) | 0.5199 (0.5026-0.5373) |
| LDpred-funct | Baseline-LD model with 75 annotations | 0.6460 (0.64-0.652) | 0.6215 (0.6155 - 0.6275) |
| DBSLMM | h2f = 0.8 | 0.5326 (0.5266-0.5386) | 0.5236 (0.5176-0.5296) |
| DBSLMM | h2f = 1 | 0.5326 (0.5266-0.5386) | 0.5236 (0.5176-0.5296) |
| DBSLMM | h2f = 1.2 | 0.5326 (0.5266-0.5386) | 0.5236 (0.5176-0.5296) |

**Table 4B.** Association of Different Polygenic Risk Scores (PRS) With Coronary Artery Disease (CAD) in Tuning Case-Control Study and in Prospective Cohort Study in Japan Biobank dataset

| | | AUC (95% CI) PRS | |
|---|---|---|---|
| **Methods** | **P-value/Tuning Parameters** | **Case-control study (N = 9,499)** | **Cohort Study (N = 7,036)** |
| Clumping and Thresholding | R2=0.02, P=0.00000005 | 0.549 (0.5454-0.5526) | 0.5341 (0.5312-0.5371) |
| Clumping and Thresholding | R2=0.02, P=0.000005 | 0.5529 (0.5494-0.5565) | 0.5395 (0.5365-0.5424) |
| Clumping and Thresholding | R2=0.02, P=0.0005 | 0.5602 (0.5567-0.5638) | 0.5488 (0.5458-0.5517) |
| Clumping and Thresholding | R2=0.02, P=0.05 | 0.5584 (0.5548-0.5619) | 0.542 (0.5391-0.545) |
| Clumping and Thresholding | R2=0.02, P=0.5 | 0.5559 (0.5523-0.5595) | 0.5401 (0.5372-0.5431) |
| Clumping and Thresholding | R2=0.02, P=1 | 0.5558 (0.5522-0.5594) | 0.5394 (0.5364-0.5423) |
| Clumping and Thresholding | R2=0.04, P=0.00000005 | 0.5492 (0.5456-0.5527) | 0.5331 (0.5301-0.536) |
| Clumping and Thresholding | R2=0.04, P=0.000005 | 0.5515 (0.5479-0.5551) | 0.5372 (0.5343-0.5402) |
| Clumping and Thresholding | R2=0.04, P=0.0005 | 0.558 (0.5545-0.5616) | 0.5472 (0.5443-0.5501) |
| Clumping and Thresholding | R2=0.04, P=0.05 | 0.567 (0.5634-0.5706) | 0.5489 (0.546-0.5519) |
| Clumping and Thresholding | R2=0.04. P=0.5 | 0.5647 (0.5611-0.5638) | 0.5448 (0.5418-0.5477) |
| Clumping and Thresholding | R2=0.04, P=1 | 0.5639 (0.5604-0.5675) | 0.5444 (0.5414-0.5477) |
| Clumping and Thresholding | R2=0.06, P=0.00000005 | 0.5471 (0.5435-0.5507) | 0.532 (0.5291-0.5349) |
| Clumping and Thresholding | R2=0.06, P=0.000005 | 0.5471 (0.5435-0.5507) | 0.532 (0.5291-0.5349) |
| Clumping and Thresholding | R2=0.06, P=0.0005 | 0.5522 (0.5486-0.5558) | 0.543 (0.54-0.5459) |
| Clumping and Thresholding | R2=0.06, P=0.05 | 0.5671 (0.5635-0.5707) | 0.5523 (0.5494-0.5553) |
| Clumping and Thresholding | R2=0.06, P=0.5 | 0.5683 (0.5647-0.5719) | 0.5488 (0.5458-0.5517) |
| Clumping and Thresholding | R2=0.06, P=1 | 0.5679 (0.5643-0.5715) | 0.548 (0.5451-0.551) |
| Clumping and Thresholding | R2=0.08, P=0.00000005 | 0.5495 (0.5459-0.5531) | 0.5339 (0.531-0.5369) |
| Clumping and Thresholding | R2=0.08, P=0.000005 | 0.5457 (0.5421-0.5493) | 0.5347 (0.5318-0.5377) |
| Clumping and Thresholding | R2=0.08, P=0.0005 | 0.551 (0.5474-0.5546) | 0.542 (0.539-0.5449) |
| Clumping and Thresholding | R2=0.08, P=0.05 | 0.567 (0.5634-0.5706) | 0.5551 (0.5522-0.5581) |
| Clumping and Thresholding | R2=0.08, P=0.5 | 0.5712 (0.5676-0.5748) | 0.552 (0.5491-0.5549) |
| Clumping and Thresholding | R2=0.08, P=1 | 0.5709 (0.5673-0.5745) | 0.5512 (0.5483-0.5542) |
| LDpred | p = 0.001 | 0.5313 (0.508-0.5543) | 0.5185 (0.4994-0.5376) |
| LDpred | p = 0.01 | 0.5327 (0.5096-0.5559) | 0.5181 (0.499-0.5372) |
| LDpred | p = 0.1 | 0.593 (0.5698-0.6161) | 0.569 (0.5499-0.5881) |

| Methods | P-value/Tuning Parameters | Case-control study (N = 9,499) | Cohort Study (N = 7,036) |
|---|---|---|---|
| LDpred | p = 1 | 0.582 (0.5589-0.6052) | 0.56 (0.5409-0.5791) |
| LDpred | p = 0.003 | 0.5311 (0.5079-0.5542) | 0.5192 (0.5001-0.5383) |
| LDpred | p = 0.03 | 0.5446 (0.5214-0.5677) | 0.5286 (0.5095-0.5477) |
| LDpred | p = 0.3 | 0.586 (0.5628-0.6091) | 0.5634 (0.5443-0.5825) |
| lassosum | s=0.2, lambda=0.001 | 0.5577 (0.5533-0.5622) | 0.5395 (0.5356-0.5434) |
| lassosum | s=0.2, lambda=0.0013 | 0.5626 (0.5582-0.5671) | 0.543 (0.5391-0.5469) |
| lassosum | s=0.2, lambda=0.0016 | 0.5688 (0.5644-0.5733) | 0.5472 (0.5433-0.5511) |
| lassosum | s=0.2, lambda=0.0021 | 0.5752 (0.5707-0.5796) | 0.5519 (0.548-0.5559) |
| lassosum | s=0.2, lambda=0.0026 | 0.5798 (0.5754-0.5843) | 0.5572 (0.5533-0.5611) |
| lassosum | s=0.2, lambda=0.0034 | 0.5837 (0.5793-0.5882) | 0.5631 (0.5592-0.567) |
| lassosum | s=0.2, lambda=0.0043 | 0.5873 (0.5829-0.5918) | 0.5685 (0.5646-0.5724) |
| lassosum | s=0.2, lambda=0.0055 | 0.5903 (0.5858-0.5947) | 0.5719 (0.5679-0.5758) |
| lassosum | s=0.2, lambda=0.007 | 0.5863 (0.5819-0.5908) | 0.5676 (0.5637-0.5715) |
| lassosum | s=0.2, lambda=0.0089 | 0.5796 (0.5752-0.5841) | 0.5596 (0.5557-0.5636) |
| lassosum | s=0.2, lambda=0.0113 | 0.5723 (0.5678-0.5767) | 0.5525 (0.5486-0.5564) |
| lassosum | s=0.2, lambda=0.0114 | 0.5656 (0.5612-0.5701) | 0.5464 (0.5424-0.5503) |
| lassosum | s=0.2, lambda=0.0183 | 0.5593 (0.5549-0.5638) | 0.5412 (0.5373-0.5451) |
| lassosum | s=0.2, lambda=0.0234 | 0.5549 (0.5505-0.5594) | 0.5387 (0.5348-0.5426) |
| lassosum | s=0.2, lambda=0.0298 | 0.5555 (0.551-0.56) | 0.54 (0.5361-0.5439) |
| lassosum | s=0.2, lambda=0.0379 | 0.5562 (0.5518-0.5607) | 0.54 (0.536-0.5439) |
| lassosum | s=0.2, lambda=0.0483 | 0.5311 (0.5266-0.5355) | 0.5191 (0.5152-0.523) |
| lassosum | s=0.2, lambda=0.0616 | 0.5311 (0.5266-0.5355) | 0.5191 (0.5152-0.523) |
| lassosum | s=0.2, lambda=0.0785 | 0.5311 (0.5266-0.5355) | 0.5191 (0.5152-0.523) |
| lassosum | s=0.2, lambda=0.1 | 0.5311 (0.5266-0.5355) | 0.5191 (0.5152-0.523) |
| lassosum | s=0.5, lambda=0.001 | 0.5684 (0.564-0.5729) | 0.5491 (0.5452-0.553) |
| lassosum | s=0.5, lambda=0.0013 | 0.5724 (0.568-0.5769) | 0.5519 (0.548-0.5558) |
| lassosum | s=0.5, lambda=0.0016 | 0.5772 (0.5728-0.5817) | 0.5552 (0.5513-0.5592) |
| lassosum | s=0.5, lambda=0.0021 | 0.5818 (0.5773-0.5862) | 0.5592 (0.5553-0.5631) |
| lassosum | s=0.5, lambda=0.0026 | 0.5853 (0.5809-0.5898) | 0.5636 (0.5597-0.5675) |
| lassosum | s=0.5, lambda=0.0034 | 0.5887 (0.5843-0.5932) | 0.569 (0.565-0.5729) |

| **Methods** | **P-value/Tuning Parameters** | **Case-control study (N = 9,499)** | **Cohort Study (N = 7,036)** |
|---|---|---|---|
| lassosum | s=0.5, lambda=0.0043 | 0.5921 (0.5876-0.5965) | 0.5736 (0.5697-0.5775) |
| lassosum | s=0.5, lambda=0.0055 | 0.594 (0.5895-0.5985) | 0.5749 (0.571-0.5788) |
| lassosum | s=0.5, lambda=0.007 | 0.5886 (0.5841-0.5931) | 0.5689 (0.565-0.5728) |
| lassosum | s=0.5, lambda=0.0089 | 0.5802 (0.5758-0.5847) | 0.5601 (0.5562-0.564) |
| lassosum | s=0.5, lambda=0.0113 | 0.5729 (0.5685-0.5774) | 0.5527 (0.5488-0.5566) |
| lassosum | s=0.5, lambda=0.0114 | 0.5663 (0.5618-0.5707) | 0.5469 (0.5429-0.5508) |
| lassosum | s=0.5, lambda=0.0183 | 0.5603 (0.5558-0.5647) | 0.5416 (0.5377-0.5455) |
| lassosum | s=0.5, lambda=0.0234 | 0.5556 (0.5512-0.5601) | 0.539 (0.5351-0.5429) |
| lassosum | s=0.5, lambda=0.0298 | 0.5566 (0.5521-0.561) | 0.5406 (0.5367-0.5446) |
| lassosum | s=0.5, lambda=0.0379 | 0.5563 (0.5518-0.5607) | 0.5399 (0.536-0.5438) |
| lassosum | s=0.5, lambda=0.0483 | 0.5311 (0.5266-0.5355) | 0.5191 (0.5152-0.523) |
| lassosum | s=0.5, lambda=0.0616 | 0.5311 (0.5266-0.5355) | 0.5191 (0.5152-0.523) |
| lassosum | s=0.5, lambda=0.0785 | 0.5311 (0.5266-0.5355) | 0.5191 (0.5152-0.523) |
| lassosum | s=0.5, lambda=0.1 | 0.5311 (0.5266-0.5355) | 0.5191 (0.5152-0.523) |
| lassosum | s=0.9, lambda=0.001 | 0.5806 (0.5761-0.585) | 0.5611 (0.5572-0.565) |
| lassosum | s=0.9, lambda=0.0013 | 0.5832 (0.5787-0.5876) | 0.5631 (0.5592-0.567) |
| lassosum | s=0.9, lambda=0.0016 | 0.5859 (0.5815-0.5904) | 0.5653 (0.5614-0.5692) |
| lassosum | s=0.9, lambda=0.0021 | 0.5884 (0.5839-0.5929) | 0.5682 (0.5642-0.5721) |
| lassosum | s=0.9, lambda=0.0026 | 0.591 (0.5866-0.5955) | 0.5716 (0.5677-0.5755) |
| lassosum | s=0.9, lambda=0.0034 | 0.5931 (0.5887-0.5976) | 0.5752 (0.5713-0.5792) |
| lassosum | s=0.9, lambda=0.0043 | 0.5944 (0.5899-0.5988) | 0.5777 (0.5738-0.5816) |
| lassosum | s=0.9, lambda=0.0055 | 0.5923 (0.5879-0.5968) | 0.5748 (0.5709-0.5787) |
| lassosum | s=0.9, lambda=0.007 | 0.5853 (0.5808-0.5898) | 0.5654 (0.5614-0.5693) |
| lassosum | s=0.9, lambda=0.0089 | 0.5771 (0.5727-0.5816) | 0.5568 (0.5528-0.5607) |
| lassosum | s=0.9, lambda=0.0113 | 0.5707 (0.5662-0.5751) | 0.5501 (0.5462-0.554) |
| lassosum | s=0.9, lambda=0.0114 | 0.5656 (0.5612-0.5701) | 0.546 (0.5421-0.5499) |
| lassosum | s=0.9, lambda=0.0183 | 0.559 (0.5545-0.5634) | 0.5405 (0.5366-0.5445) |
| lassosum | s=0.9, lambda=0.0234 | 0.5558 (0.5513-0.5602) | 0.5384 (0.5345-0.5423) |
| lassosum | s=0.9, lambda=0.0298 | 0.5578 (0.5533-0.5623) | 0.5405 (0.5366-0.5444) |
| lassosum | s=0.9, lambda=0.0379 | 0.5563 (0.5518-0.5608) | 0.5398 (0.5359-0.5438) |

| Methods | P-value/Tuning Parameters | Case-control study (N = 9,499) | Cohort Study (N = 7,036) |
|---|---|---|---|
| lassosum | s=0.9, lambda=0.0483 | 0.5311 (0.5266-0.5355) | 0.5191 (0.5152-0.523) |
| lassosum | s=0.9, lambda=0.0616 | 0.5311 (0.5266-0.5355) | 0.5191 (0.5152-0.523) |
| lassosum | s=0.9, lambda=0.0785 | 0.5311 (0.5266-0.5355) | 0.5191 (0.5152-0.523) |
| lassosum | s=0.9, lambda=0.1 | 0.5311 (0.5266-0.5355) | 0.5191 (0.5152-0.523) |
| lassosum | s=1, lambda=0.001 | 0.5723 (0.5678-0.5767) | 0.5572 (0.5533-0.5612) |
| lassosum | s=1, lambda=0.0013 | 0.5723 (0.5678-0.5768) | 0.5576 (0.5537-0.5615) |
| lassosum | s=1, lambda=0.0016 | 0.5721 (0.5676-0.5765) | 0.5579 (0.554-0.5618) |
| lassosum | s=1, lambda=0.0021 | 0.5713 (0.5669-0.5758) | 0.5578 (0.5539-0.5617) |
| lassosum | s=1, lambda=0.0026 | 0.5697 (0.5653-0.5742) | 0.5569 (0.5529-0.5608) |
| lassosum | s=1, lambda=0.0034 | 0.5672 (0.5628-0.5717) | 0.5548 (0.5508-0.5587) |
| lassosum | s=1, lambda=0.0043 | 0.5639 (0.5595-0.5684) | 0.5511 (0.5472-0.5551) |
| lassosum | s=1, lambda=0.0055 | 0.5599 (0.5554-0.5643) | 0.5464 (0.5425-0.5503) |
| lassosum | s=1, lambda=0.007 | 0.557 (0.5525-0.5614) | 0.5421 (0.5382-0.546) |
| lassosum | s=1, lambda=0.0089 | 0.5559 (0.5515-0.5604) | 0.5392 (0.5353-0.5431) |
| lassosum | s=1, lambda=0.0113 | 0.5574 (0.5529-0.5618) | 0.5379 (0.534-0.5418) |
| lassosum | s=1, lambda=0.0114 | 0.5577 (0.5532-0.5621) | 0.5373 (0.5333-0.5412) |
| lassosum | s=1, lambda=0.0183 | 0.5552 (0.5507-0.5596) | 0.5374 (0.5335-0.5413) |
| lassosum | s=1, lambda=0.0234 | 0.5513 (0.5469-0.5558) | 0.5343 (0.5304-0.5383) |
| lassosum | s=1, lambda=0.0298 | 0.5553 (0.5509-0.5598) | 0.5379 (0.534-0.5418) |
| lassosum | s=1, lambda=0.0379 | 0.5566 (0.5521-0.5611) | 0.54 (0.5361-0.544) |
| lassosum | s=1, lambda=0.0483 | 0.5311 (0.5266-0.5355) | 0.5191 (0.5152-0.523) |
| lassosum | s=1, lambda=0.0616 | 0.5311 (0.5266-0.5355) | 0.5191 (0.5152-0.523) |
| lassosum | s=1, lambda=0.0785 | 0.5311 (0.5266-0.5355) | 0.5191 (0.5152-0.523) |
| lassosum | s=1, lambda=0.1 | 0.5311 (0.5266-0.5355) | 0.5191 (0.5152-0.523) |
| PRS-CS | phi = 0.000001 | 0.583 (0.5619-0.604) | 0.5602 (0.5415-0.5788) |
| PRS-CS | phi =0.0001 | 0.597 (0.576-0.6181) | 0.5752 (0.5565-0.5939) |
| PRS-CS | phi = 0.01 | 0.5811 (0.5601-0.6022) | 0.5589 (0.5403-0.5776) |
| PRS-CS | phi = 1 | 0.5647 (0.5436-0.5858) | 0.5466 (0.5279-0.5652) |
| sBayesR | LD matrix = HapMap3 and 2.8M | 0.5308 (0.5248-0.5368) | 0.5186 (0.5126-0.5246) |
| sBayesR | LD matrix = 2.8M variants | 0.5328 (0.5268-0.5388) | 0.5177 (0.5117-0.5237) |

| Methods | P-value/Tuning Parameters | Case-control study (N = 9,499) | Cohort Study (N = 7,036) |
|---|---|---|---|
| LDpred-funct | Baseline-LD model with 75 annotations | 0.5907 (0.5847-0.5967) | 0.5692 (0.5632-0.5752) |
| DBSLMM | h2f = 0.8 | 0.5347 (0.5314-0.5381) | 0.5258 (0.5201-0.5314) |
| DBSLMM | h2f = 1 | 0.5353 (0.5319-0.5386) | 0.5263 (0.5207-0.532) |
| DBSLMM | h2f = 1.2 | 0.5358 (0.5324-0.5391) | 0.5268 (0.5212-0.5324) |

**Table 5A.** C Statistics (Derived for Cox Regression) for CAD Using Recalibrated Models in the PCE Prospective Cohort, Primary Analysis. Results are shown for the European meta-analysis and Japan Biobank datasets. Results are presented for the full population and stratified by gender and age group (below or above 55 years of age). Results for the sensitivity analysis using only participants with no reported lipid-lowering treatment at baseline also shown.

A. European Meta-Analysis

| | All Participants (N=272,307; 7,036 cases) | Men (N=124,155; 5093 cases) | Women (N=148,152; 1,943 cases) | < 55 years old (N=102,330; 1,276 cases) | ≥ 55 years old (N=169,977; 5,760 cases) | Participants Not Receiving Lipid-Lowering Treatment at Baseline (N= 235,172; 5,091 cases) |
|---|---|---|---|---|---|---|
| **PRS** | 0.636 (0.63-0.642) | 0.639 (0.632-0.647) | 0.638 (0.625-0.65) | 0.683 (0.669-0.698) | 0.629 (0.622-0.636) | 0.642 (0.634-0.649) |
| **PCE** | 0.718 (0.713-0.723) | 0.663 (0.656-0.67) | 0.706 (0.695-0.717) | 0.749 (0.736-0.761) | 0.665 (0.658-0.671) | 0.73 (0.724-0.737) |
| **PRS + PCE** | 0.752 (0.746-0.757) | 0.712 (0.705-0.718) | 0.74 (0.729-0.75) | 0.791 (0.779-0.803) | 0.704 (0.698-0.71) | 0.764 (0.758-0.77) |

B. Japan Biobank Dataset

| | All Participants (N=272,307; 7,036 cases) | Men (N=124,155; 5093 cases) | Women (N=148,152; 1,943 cases) | < 55 years old (N=102,330; 1,276 cases) | ≥ 55 years old (N=169,977; 5,760 cases) | Participants Not Receiving Lipid-Lowering Treatment at Baseline (N= 235,172; 5,091 cases) |
|---|---|---|---|---|---|---|
| **PRS** | 0.582 (0.576-0.589) | 0.582 (0.575-0.59) | 0.584 (0.572-0.597) | 0.625 (0.61-0.64) | 0.576 (0.568-0.583) | 0.583 (0.575-0.591) |
| **PCE** | 0.718 (0.713-0.723) | 0.663 (0.656-0.67) | 0.706 (0.695-0.717) | 0.749 (0.736-0.761) | 0.665 (0.658-0.671) | 0.73 (0.724-0.737) |
| **PRS + PCE** | 0.73 (0.725-0.735) | 0.681 (0.675-0.688) | 0.718 (0.707-0.729) | 0.767 (0.755-0.78) | 0.678 (0.671-0.684) | 0.743 (0.737-0.749) |

**Table 5B.** C Statistics (Derived from Cox Regression) for CAD Using European Meta-analysis dataset and Recalibrated Models in the PCE Prospective Cohort, Primary Analysis. Results are presented for full population and stratified by gender and age group (below or above 55 years of age). Results for the sensitivity analysis using only participants with no reported lipid-lowering treatment at baseline also shown.

| A. Full Population (N=272,307; 7036 cases) | | | | | | | |
|---|---|---|---|---|---|---|---|
| | **Clumping and Thresholding** | **LDpred** | **lassosum** | **PRS-CS** | **sBayesR** | **LDpred-funct** | **DBSLMM** |
| **PRS** | 0.603 (0.597-0.61) | 0.63 (0.624-0.636) | 0.628 (0.621-0.634) | 0.63 (0.624-0.636) | 0.603 (0.597-0.61) | 0.62 (0.614-0.627) | 0.523 (0.517-0.53) |
| **PCE** | 0.718 (0.713-0.723) | 0.718 (0.713-0.723) | 0.718 (0.713-0.723) | 0.718 (0.713-0.723) | 0.718 (0.713-0.723) | 0.718 (0.713-0.723) | 0.718 (0.713-0.723) |
| **PRS + PCE** | 0.738 (0.732-0.743) | 0.749 (0.743-0.754) | 0.748 (0.743-0.753) | 0.749 (0.744-0.754) | 0.738 (0.732-0.743) | 0.743 (0.738-0.749) | 0.718 (0.713-0.724) |
| B. Men (N=124,155; 5093 cases) | | | | | | | |
| | **Clumping and Thresholding** | **LDpred** | **lassosum** | **PRS-CS** | **sBayesR** | **LDpred-funct** | **DBSLMM** |
| **PRS** | 0.606 (0.599-0.614) | 0.632 (0.624-0.639) | 0.63 (0.623-0.638) | 0.633 (0.625-0.64) | 0.606 (0.599-0.614) | 0.621 (0.613-0.628) | 0.515 (0.507-0.523) |
| **PCE** | 0.663 (0.656-0.67) | 0.663 (0.656-0.67) | 0.663 (0.656-0.67) | 0.663 (0.656-0.67) | 0.663 (0.656-0.67) | 0.663 (0.656-0.67) | 0.663 (0.656-0.67) |
| **PRS + PCE** | 0.688 (0.681-0.695) | 0.703 (0.697-0.71) | 0.703 (0.697-0.71) | 0.704 (0.697-0.711) | 0.688 (0.681-0.695) | 0.696 (0.69-0.703) | 0.661 (0.654-0.668) |
| C. Women (N=148,152; 1943 cases) | | | | | | | |
| | **Clumping and Thresholding** | **LDpred** | **lassosum** | **PRS-CS** | **sBayesR** | **LDpred-funct** | **DBSLMM** |
| **PRS** | 0.602 (0.589-0.614) | 0.632 (0.619-0.644) | 0.622 (0.609-0.635) | 0.629 (0.617-0.642) | 0.602 (0.589-0.614) | 0.622 (0.61-0.635) | 0.527 (0.514-0.54) |
| **PCE** | 0.706 (0.695-0.717) | 0.706 (0.695-0.717) | 0.706 (0.695-0.717) | 0.706 (0.695-0.717) | 0.706 (0.695-0.717) | 0.706 (0.695-0.717) | 0.706 (0.695-0.717) |
| **PRS + PCE** | 0.725 (0.714-0.735) | 0.736 (0.726-0.747) | 0.732 (0.722-0.743) | 0.736 (0.726-0.747) | 0.725 (0.714-0.735) | 0.731 (0.72-0.742) | 0.705 (0.695-0.716) |
| D. Aged < 55 years old (N=102,330; 1276 cases) | | | | | | | |
| | **Clumping and Thresholding** | **LDpred** | **lassosum** | **PRS-CS** | **sBayesR** | **LDpred-funct** | **DBSLMM** |
| **PRS** | 0.64 (0.625-0.656) | 0.675 (0.66-0.69) | 0.67 (0.655-0.685) | 0.675 (0.66-0.69) | 0.64 (0.625-0.656) | 0.662 (0.647-0.677) | 0.541 (0.525-0.557) |
| **PCE** | 0.749 (0.736-0.761) | 0.749 (0.736-0.761) | 0.749 (0.736-0.761) | 0.749 (0.736-0.761) | 0.749 (0.736-0.761) | 0.749 (0.736-0.761) | 0.749 (0.736-0.761) |
| **PRS + PCE** | 0.77 (0.757-0.782) | 0.783 (0.771-0.795) | 0.783 (0.77-0.795) | 0.784 (0.772-0.796) | 0.77 (0.757-0.782) | 0.779 (0.767-0.792) | 0.747 (0.734-0.76) |
| E. Aged ≥ 55 years old (N=169,977; 5,760 cases) | | | | | | | |
| | **Clumping and Thresholding** | **LDpred** | **lassosum** | **PRS-CS** | **sBayesR** | **LDpred-funct** | **DBSLMM** |
| **PRS** | 0.599 (0.592-0.606) | 0.623 (0.616-0.63) | 0.621 (0.614-0.629) | 0.623 (0.616-0.63) | 0.599 (0.592-0.606) | 0.614 (0.607-0.621) | 0.522 (0.515-0.53) |
| **PCE** | 0.665 (0.658-0.671) | 0.665 (0.658-0.671) | 0.665 (0.658-0.671) | 0.665 (0.658-0.671) | 0.665 (0.658-0.671) | 0.665 (0.658-0.671) | 0.665 (0.658-0.671) |
| **PRS + PCE** | 0.685 (0.679-0.692) | 0.698 (0.692-0.704) | 0.697 (0.691-0.703) | 0.698 (0.692-0.705) | 0.685 (0.679-0.692) | 0.692 (0.685-0.698) | 0.664 (0.658-0.671) |
| F. Participants not receiving lipid-lowering treatment at baseline (N=235,172; 5,091 cases) | | | | | | | |
| | **Clumping and Thresholding** | **LDpred** | **lassosum** | **PRS-CS** | **sBayesR** | **LDpred-funct** | **DBSLMM** |
| **PRS** | 0.598 (0.59-0.606) | 0.629 (0.621-0.636) | 0.627 (0.619-0.634) | 0.63 (0.622-0.637) | 0.517 (0.509-0.525) | 0.623 (0.616-0.631) | 0.521 (0.513-0.529) |
| **PCE** | 0.73 (0.724-0.737) | 0.73 (0.724-0.737) | 0.73 (0.724-0.737) | 0.73 (0.724-0.737) | 0.73 (0.724-0.737) | 0.73 (0.724-0.737) | 0.73 (0.724-0.737) |
| **PRS + PCE** | 0.746 (0.74-0.752) | 0.758 (0.752-0.764) | 0.757 (0.751-0.763) | 0.759 (0.753-0.765) | 0.73 (0.724-0.736) | 0.755 (0.749-0.761) | 0.73 (0.723-0.736) |

**Table 5C.** C Statistics (Derived from Cox Regression) for CAD Using Japan Biobank dataset and Recalibrated Models in the PCE Prospective Cohort, Primary Analysis. Results are presented for full population and stratified by gender and age group (below or above 55 years of age). Results for the sensitivity analysis using only participants with no reported lipid-lowering treatment at baseline also shown.

| A. Full Population (N=272,307; 7036 cases) | | | | | | | |
|---|---|---|---|---|---|---|---|
| | **Clumping and Thresholding** | **LDpred** | **lassosum** | **PRS-CS** | **sBayesR** | **LDpred-funct** | **DBSLMM** |
| **PRS** | 0.552 (0.545-0.558) | 0.568 (0.562-0.575) | 0.577 (0.57-0.583) | 0.574 (0.568-0.581) | 0.552 (0.545-0.558) | 0.569 (0.562-0.575) | 0.527 (0.52-0.533) |
| **PCE** | 0.718 (0.713-0723) | 0.718 (0.713-0723) | 0.718 (0.713-0723) | 0.718 (0.713-0723) | 0.718 (0.713-0723) | 0.718 (0.713-0723) | 0.718 (0.713-0723) |
| **PRS + PCE** | 0.722 (0.717-0.727) | 0.726 (0.72-0.731) | 0.727 (0.721-0.732) | 0.727 (0.722-0.733) | 0.722 (0.717-0.727) | 0.726 (0.721-0.732) | 0.718 (0.713-0724) |
| B. Men (N=124,155; 5093 cases) | | | | | | | |
| | **Clumping and Thresholding** | **LDpred** | **lassosum** | **PRS-CS** | **sBayesR** | **LDpred-funct** | **DBSLMM** |
| **PRS** | 0.548 (0.54-0.556) | 0.566 (0.558-0.574) | 0.578 (0.571-0.586) | 0.573 (0.566-0.581) | 0.548 (0.54-0.556) | 0.565 (0.558-0.573) | 0.519 (0.511-0.527) |
| **PCE** | 0.663 (0.656-0.67) | 0.663 (0.656-0.67) | 0.663 (0.656-0.67) | 0.663 (0.656-0.67) | 0.663 (0.656-0.67) | 0.663 (0.656-0.67) | 0.663 (0.656-0.67) |
| **PRS + PCE** | 0.666 (0.659-0.673) | 0.671 (0.664-0.678) | 0.675 (0.668-0.682) | 0.674 (0.667-0.681) | 0.666 (0.659-0.673) | 0.671 (0.665-0.678) | 0.66 (0.653-0.667) |
| C. Women (N=148,152; 1943 cases) | | | | | | | |
| | **Clumping and Thresholding** | **LDpred** | **lassosum** | **PRS-CS** | **sBayesR** | **LDpred-funct** | **DBSLMM** |
| **PRS** | 0.553 (0.54-0.566) | 0.571 (0.559-0.584) | 0.571 (0.559-0.584) | 0.575 (0.562-0.588) | 0.553 (0.54-0.566) | 0.574 (0.562-0.587) | 0.524 (0.511-0.538) |
| **PCE** | 0.706 (0.695-0.717) | 0.706 (0.695-0.717) | 0.706 (0.695-0.717) | 0.706 (0.695-0.717) | 0.706 (0.695-0.717) | 0.706 (0.695-0.717) | 0.706 (0.695-0.717) |
| **PRS + PCE** | 0.707 (0.697-0.718) | 0.713 (0.702-0.723) | 0.712 (0.701-0.723) | 0.714 (0.703-0.724) | 0.707 (0.697-0.718) | 0.714 (0.704-0.725) | 0.705 (0.694-0.716) |
| D. Aged < 55 years old (N=102,330; 1276 cases) | | | | | | | |
| | **Clumping and Thresholding** | **LDpred** | **lassosum** | **PRS-CS** | **sBayesR** | **LDpred-funct** | **DBSLMM** |
| **PRS** | 0.586 (0.571-0.602) | 0.605 (0.589-0.62) | 0.618 (0.603-0.633) | 0.616 (0.601-0.632) | 0.586 (0.571-0.602) | 0.606 (0.591-0.622) | 0.55 (0.534-0.566) |
| **PCE** | 0.749 (0.736-0.761) | 0.749 (0.736-0.761) | 0.749 (0.736-0.761) | 0.749 (0.736-0.761) | 0.749 (0.736-0.761) | 0.749 (0.736-0.761) | 0.749 (0.736-0.761) |
| **PRS + PCE** | 0.755 (0.742-0.768) | 0.758 (0.745-0.77) | 0.762 (0.75-0.775) | 0.762 (0.75-0.775) | 0.755 (0.742-0.768) | 0.759 (0.747-0.772) | 0.748 (0.735-0.76) |
| E. Aged ≥ 55 years old (N=169,977; 5,760 cases) | | | | | | | |
| | **Clumping and Thresholding** | **LDpred** | **lassosum** | **PRS-CS** | **sBayesR** | **LDpred-funct** | **DBSLMM** |
| **PRS** | 0.544 (0.537-0.552) | 0.562 (0.555-0.569) | 0.567 (0.56-0.574) | 0.567 (0.56-0.574) | 0.544 (0.537-0.552) | 0.562 (0.554-0.569) | 0.526 (0.518-0.533) |
| **PCE** | 0.665 (0.658-0.671) | 0.665 (0.658-0.671) | 0.665 (0.658-0.671) | 0.665 (0.658-0.671) | 0.665 (0.658-0.671) | 0.665 (0.658-0.671) | 0.665 (0.658-0.671) |
| **PRS + PCE** | 0.667 (0.66-0.674) | 0.671 (0.664-0.678) | 0.672 (0.666-0.679) | 0.673 (0.666-0.679) | 0.667 (0.66-0.674) | 0.672 (0.665-0.678) | 0.664 (0.658-0.671) |
| F. Participants not receiving lipid-lowering treatment at baseline (N=235,172; 5,091 cases) | | | | | | | |
| | **Clumping and Thresholding** | **LDpred** | **lassosum** | **PRS-CS** | **sBayesR** | **LDpred-funct** | **DBSLMM** |
| **PRS** | 0.549 (0.542-0.557) | 0.566 (0.559-0.574) | 0.569 (0.562-0.577) | 0.572 (0.564-0.58) | 0.515 (0.507-0.523) | 0.566 (0.558-0.574) | 0.525 (0.517-0.533) |
| **PCE** | 0.73 (0.724-0.737) | 0.73 (0.724-0.737) | 0.73 (0.724-0.737) | 0.73 (0.724-0.737) | 0.73 (0.724-0.737) | 0.73 (0.724-0.737) | 0.73 (0.724-0.737) |
| **PRS + PCE** | 0.734 (0.727-0.74) | 0.738 (0.731-0.744) | 0.739 (0.733-0.745) | 0.739 (0.733-0.745) | 0.729 (0.723-0.735) | 0.738 (0.732-0.744) | 0.73 (0.724-0.736) |

**Table 5D.** C Statistics (Derived for Cox Regression) for CAD Using Recalibrated Models in the PCE Prospective Cohort, Secondary Analysis. Results are shown for the European meta-analysis and Japan Biobank datasets. Results are presented for the full population and stratified by gender and age group (below or above 55 years of age). Results for the sensitivity analysis using only participants with no reported lipid-lowering treatment at baseline also shown.

A. European Meta-Analysis

| | **All Participants (N= 6,753; 88 cases)** | **Men (N= 2,901; 46 cases)** | **Women (N= 3,852; 42 cases)** | **< 55 years old (N=4,528; 42 cases)** | **≥ 55 years old (N=2,225; 46 cases)** | **Participants Not Receiving Lipid-Lowering Treatment at Baseline (N= 5,896; 63 cases)** |
|---|---|---|---|---|---|---|
| **PRS** | 0.545 (0.488-0.564) | 0.572 (0.49-0.653) | 0.6 (0.512-0.688) | 0.549 (0.461-0.637) | 0.551 (0.472-0.63) | 0.54 (0.471-0.609) |
| **PCE** | 0.714 (0.659-0.769) | 0.674 (0.595-0.753) | 0.734 (0.653-0.815) | 0.657 (0.572-0.742) | 0.721 (0.656-0.787) | 0.698 (0.628-0.768) |
| **PRS + PCE** | 0.715 (0.662–0.768) | 0.708 (0.636-0.78) | 0.734 (0.657-0.811) | 0.671 (0.587-0.755) | 0.7 (0.634-0.767) | 0.71 (0.643-0.777) |

B. Japan Biobank Dataset

| | **All Participants (N= 6,753; 88 cases)** | **Men (N= 2,901; 46 cases)** | **Women (N= 3,852; 42 cases)** | **< 55 years old (N=4,528; 42 cases)** | **≥ 55 years old (N=2,225; 46 cases)** | **Participants Not Receiving Lipid-Lowering Treatment at Baseline (N= 5,896; 63 cases)** |
|---|---|---|---|---|---|---|
| **PRS** | 0.533 (0.472-0.595) | 0.497 (0.409-0.585) | 0.549 (0.463-0.634) | 0.551 (0.463-0.639) | 0.506 (0.417-0.595) | 0.522 (0.452-0.591) |
| **PCE** | 0.714 (0.659-0.769) | 0.674 (0.595-0.753) | 0.734 (0.653-0.815) | 0.657 (0.572-0.742) | 0.721 (0.656-0.787) | 0.698 (0.628-0.768) |
| **PRS + PCE** | 0.714 (0.66-0.769) | 0.668 (0.589-0.748) | 0.735 (0.653-0.816) | 0.666 (0.582-0.75) | 0.719 (0.653-0.785) | 0.697 (0.627-0.767) |

**Table 6.** Risk Reclassification at 7.5% Threshold for CAD Using Recalibrated PCE and PCE + PRS Models Stratified by Gender and Age Group (below or above 55 years of age)

| | | | | | | | |
|---|---|---|---|---|---|---|---|
| **Men** | | | | | | | |
| | | | PCE + PRS | | | | |
| | | | < 7.5% | ≥ 7.5% | % reclassified | Categorical NRI: | Continuous NRI: |
| PCE | < 7.5% | Cases | 2881 | 1129 | 22.3 | 0.1671 (0.1532 to 0.1762) | 0.238 (0.2131 to 0.2599) |
| | ≥ 7.5% | Cases | 281 | 783 | 5.5 | | |
| | < 7.5% | Noncases | 92944 | 10405 | 9.1 | -0.051 (-0.056 to -0.047) | 0.1722 (0.1634 to 0.1849) |
| | ≥ 7.5% | Noncases | 4517 | 5894 | 4.0 | | |
| | | | | | NRI in full population: | 0.1161 (0.1046 to 0.124) | 0.4102 (0.377 to 0.4448) |
| | IDI: | 0.0708 (0.0674 to 0.0743) | | | | | |
| **Women** | | | | | | | |
| | | | PCE + PRS | | | | |
| | | | < 7.5% | ≥ 7.5% | % reclassified | Categorical NRI: | Continuous NRI: |
| PCE | < 7.5% | Cases | 1803 | 88 | 4.6 | 0.0413 (0.0302 to 0.0497) | 0.0962 (0.0837 to 0.1349) |
| | ≥ 7.5% | Cases | 10 | 30 | 0.5 | | |
| | < 7.5% | Noncases | 140610 | 999 | 0.7 | -0.0056 (-0.0066 to -0.0045) | 0.3196 (0.3108 to 0.3611) |
| | ≥ 7.5% | Noncases | 244 | 459 | 0.2 | | |
| | | | | | NRI in full population: | 0.0357 (0.0248 to 0.0431) | 0.4158 (0.4057 to 0.496) |
| | IDI: | 0.0452 (0.0398 to 0.0507) | | | | | |
| **Aged < 55 years old** | | | | | | | |
| | | | PCE + PRS | | | | |
| | | | < 7.5% | ≥ 7.5% | % reclassified | Categorical NRI: | Continuous NRI: |
| PCE | < 7.5% | Cases | 1053 | 140 | 11.0 | 0.0889 (0.0701 to 0.0984) | 0.1683 (0.1063 to 0.2012) |
| | ≥ 7.5% | Cases | 28 | 46 | 2.2 | | |
| | < 7.5% | Noncases | 97435 | 1244 | 1.2 | -0.0076 (-0.0088 to -0.0066) | 0.3438 (0.2509 to 0.4) |
| | ≥ 7.5% | Noncases | 497 | 401 | 0.5 | | |
| | | | | | NRI in full population: | 0.0813 (0.0626 to 0.0901) | 0.5121 (0.3776 to 0.6012) |
| | IDI: | 0.0702 (0.0628 to 0.0776) | | | | | |
| **Aged ≥ 55 years old** | | | | | | | |
| | | | PCE + PRS | | | | |
| | | | < 7.5% | ≥ 7.5% | % reclassified | Categorical NRI: | Continuous NRI: |
| PCE | < 7.5% | Cases | 4093 | 902 | 15.7 | 0.123 (0.1192 to 0.1336) | 0.1897 (0.1698 to 0.2285) |
| | ≥ 7.5% | Cases | 200 | 543 | 3.5 | | |
| | < 7.5% | Noncases | 140310 | 8769 | 5.6 | -0.0359 (-0.0391 to -0.0344) | 0.1617 (0.1569 to 0.1827) |
| | ≥ 7.5% | Noncases | 3163 | 4253 | 2.0 | | |
| | | | | | NRI in full population: | 0.087 (0.0843 to 0.0969) | 0.3514 (0.3304 to 0.4112) |
| | IDI: | 0.0605 (0.0572 to 0.0639) | | | | | |

**Table 7.** Net Reclassification Improvement and Integrated Discrimination Improvement Results in Secondary Analysis of 6,753 African Ancestry Participants

| | No. of Participants | Continuous Net Reclassification Improvement | Categorical Net Reclassification Improvement | Integrated Discrimination Improvement |
|---|---|---|---|---|
| Cases | 88 | 0.044 (0.036 to 0.052) | 0.0 (-.0.46 to 0.0) | |
| Noncases | 6514 | -0.046 (-0.054 to -0.038) | 0.0 (0.0 to 0.005) | |
| Full Population | 6602 | -0.002 (-0.203 to 0.198) | 0.0 (-0.041 to 0.001) | 0.0129 (-0.0076 to -0.0333) |
| Censored | 151 | | | |

**Figure 1.** Correlation of PRSs Constructed by Different Methods in the Tuning Set of 9,499 Prevalent CAD cases and an Equal Number of Controls.
Abbreviations: PT and LDpredfun refer to the Clumping and Thresholding and LDpred-funct methods, respectively.

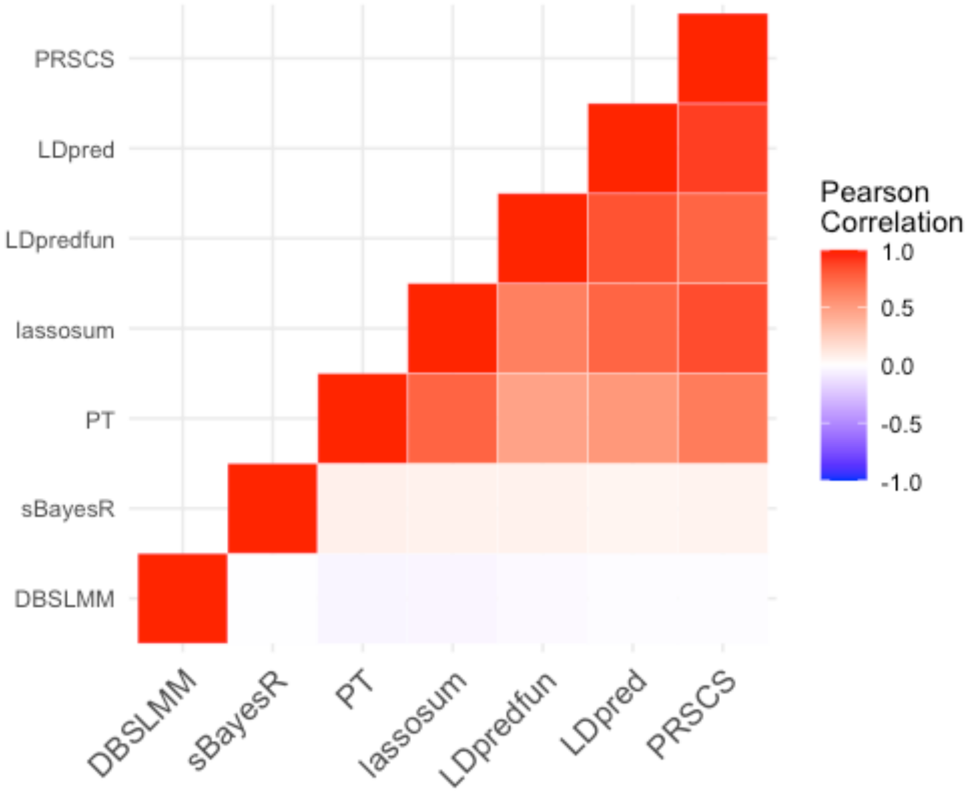

**Figure 2.** Calibration and Recalibration Plots Polygenic Risk Score for Coronary Artery Disease (CAD) and PRS Combined with PCE, Using a UK Biobank Prospective Cohort Sample. Model performance shown for non-recalibrated and recalibrated models. Recalibration was performed by fitting the predicted log hazard of the original models as a covariate in a Cox survival model. $P_{GND}$ is the associated Greenwood-Nam-D'Agostino test P-value that tests a null hypothesis of the observed and expected probabilities being identical in each group.

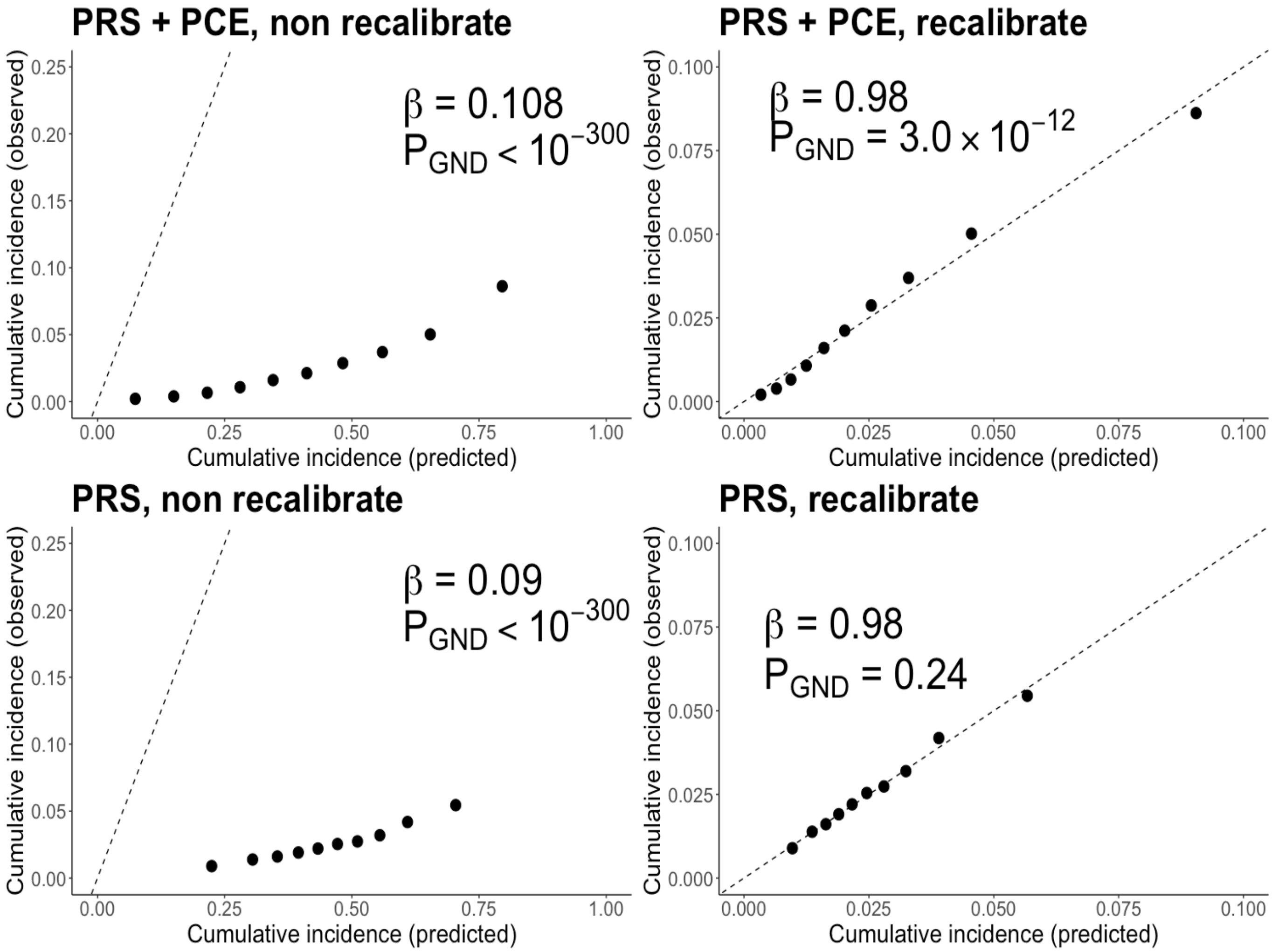
